# Estimating malaria antigen dynamics and the time to negativity of next-generation malaria rapid diagnostic tests

**DOI:** 10.1101/2025.01.13.25320483

**Authors:** William Sheahan, Allison Golden, Rebecca Barney, Smita Das, Ihn-Kyung Jang, Henry Ntuku, Xue Wu, Brooke Whittemore, Lucille Dausab, Davis Mumbengegwi, Hannah Slater, Gonzalo J Domingo, Michelle S Hsiang

## Abstract

**Background:** Rapid diagnostic tests (RDTs) used to diagnose *Plasmodium falciparum* (Pf) predominantly target the antigen Histidine Rich Protein 2 (HRP2) exclusively. With the emergence of *hrp2/hrp3* gene deletions, RDTs targeting other antigens such as the essential enzyme Lactate Dehydrogenase (LDH) are needed. The dynamics of LDH relative to HRP2 are currently not well described but are needed to inform the use of next-generation (NG-) LDH and HRP2 RDTs that are designed to address *hrp2/hrp3* gene deletions.

**Methods:** A longitudinal cohort study conducted in a low transmission setting in Namibia was leveraged to compare HRP2 and LDH decay rates. Passive and active case detection were used to recruit individuals with positive HRP2-RDT results. Study participants were treated and subsequently followed weekly until they received two consecutive HRP2-RDT negative results. Blood specimens were characterized for antigen concentration and parasite density. Antigen decay rates were calculated and used to estimate time to negativity (TTN) of NG-RDTs: two HRP2 and LDH-based RDTs (Rapigen Pf and a WHO prequalified Pf/Pv RDT) and an LDH-only RDT (Rapigen Pf/Pv).

**Results:** In 135 participants, the starting geometric mean concentrations for HRP2 and LDH were 899 ng/mL and 344 ng/mL respectively. Both antigens followed a biphasic decay rate, with a faster decay rate in the first phase. For current RDTs with an analytical sensitivity of 1 ng/mL for HRP2 and 5 ng/mL for LDH, TTN was 44 and 4 days, respectively. With a NG-RDT with LDH analytical sensitivity of 0.37 ng/mL, average TTN was 9 days. Multiple levels of analytical sensitivity were also modeled.

**Conclusions:** In the detection of Pf malaria, LDH versus HRP2-based RDTs had a faster TTN due to a combination of lower accumulated antigen concentrations and faster decay rates, even for more sensitive LDH-based RDTs. Detection of LDH versus HRP2 by RDT is more likely to reflect a new or very recent infection. For NG-RDTs that target both antigens, HRP2 is likely to contribute more to the test signal than LDH in recently treated infections unless the infection has *hrp2/hrp3* gene deletions. Antigen decay data combined with analytical sensitivity of RDTs contributes to our understanding of their performance and interpretation.

## Introduction

Key to the control of malaria is accurate diagnosis of infection before initiating antimalarial treatment, as recommended by the World Health Organization (WHO).^1^ In the Republic of Namibia (hereafter Namibia), national policy aligns with WHO in recommending that suspected cases presenting at health facilities be confirmed by rapid diagnostic test (RDT) or microscopy before treatment.^2^ In 2021, 99.9 percent of all malaria diagnostics conducted in Namibia were RDTs,^3^ and Access Bio’s *CareStart*™ Malaria Pf/PAN (HRP2/pLDH) Antigen Combo RDT was the standard RDT used at the time of this study (hereafter Standard RDT).^4^

Malaria RDTs detect antigens produced by malarial parasites, either the histidine-rich protein 2 (HRP2), which is expressed by *Plasmodium falciparum* (Pf), or *Plasmodium* lactate dehydrogenase (pLDH), which is expressed by all *Plasmodium* spp. parasites. HRP2-based RDTs are most commonly used for the diagnosis of *P. falciparum*,^5^ in part due to their more consistent performance in analytical sensitivity when compared to pLDH-based RDTs.^6^ HRP2-based RDTs have demonstrated excellent diagnostic sensitivity for *P. falciparum* infection at parasite densities of greater than 200 parasites per microliter^7,8^ but may not perform as well in low-transmission settings where parasitemia is often below the limit of detection (LOD) of commercially available RDTs.^9^

RDTs with improved analytical sensitivity (lower LOD) for malaria antigens are becoming available, including Abbott’s NxTek™ Eliminate Malaria Pf ultrasensitive rapid diagnostic test (hereafter uRDT),^10^ which has a tenfold lower LOD^11^ for HRP2 and is shown to consistently perform better in detecting asymptomatically infected individuals compared to standard RDTs.^12,13,14^ Rapigen also has improved-sensitivity RDTs on the market,^15^ which have enhanced analytical sensitivity for HRP2 as well as pLDH in comparison to most currently available RDTs.^16^ RDTs with an improved sensitivity for *P.falciparum* LDH are particularly relevant in context of the growing prevalence of *P.falciparum* infections evading detection of HRP2-based RDTs through deletion of the genes expressing HRP2 and HRP3 (a highly homologous protein that may cross react with HRP2 antibodies).^17,18^ The WHO considers *hrp2/hrp3* gene deletions one of four biological threats to malaria control and elimination.^19^ In response to this threat, WHO has published a response plan to *hrp2/hrp3* deletions^20^ as well as recommendations for procurement of LDH-based tests with higher sensitivity in settings where the prevalence of false negatives arising from hrp2/hrp3 deletions is greater than 5%. A recent study in Djibouti, where > 50% of infections may be missed by HRP2-based RDTs, showed the Rapigen tests with improved LOD for LDH to be more sensitive in febrile malaria case diagnosis than HRP2-based tests, but still not ideal.^21^ Modelling of the performance of RDTs on data from clinical specimens with associated HRP2 and LDH concentrations suggests that next generation RDTs which detect both HRP2 and pfLDH on separate lines or on a single line will perform better across more settings, and importantly will do so regardless of underlying hrp2/hrp3 deletion rates, than high sensitivity RDTs based solely on either HRP2 or LDH.^16^

The posttreatment time to negativity (TTN) of an RDT is based on the decay rate of the target antigen, the LOD of the test, and the initial antigen concentrations in individuals receiving treatment. Typically, HRP2 decays more slowly than pLDH, meaning HRP2-based RDTs will remain positive longer after treatment.^22^ Using data from studies where individuals have been treated and then tested with an RDT at regular intervals posttreatment, it is possible to estimate the TTN for different RDTs. In many cases, this type of study has not yet been conducted for new RDTs outside of the context of highly controlled clinical trials.^11,23,24^ However, by having both analytical estimates of the LODs of new RDTs and data on antigen dynamics in treated populations, it is possible to estimate the TTN of these new RDTs computationally in clinical populations.

Quantifying antigen decay and TTN is particularly important to understand the likelihood of false-positive tests among recently treated febrile patients in a clinical setting. Additionally, estimating the TTN of new and existing RDTs in recently treated populations provides potential insights into the appropriate use of RDTs in test-and-treat campaigns.

In this study, data from a longitudinal posttreatment study in Namibia are used to estimate the posttreatment dynamics of pLDH for the detection of *P. falciparum* malaria, complementing the HRP2 results first reported by Ntuku *et al.*, 2024.^4^ Additionally, using laboratory-generated benchmarking data on the LOD of next-generation RDTs^16^ with lower limits of detection for LDH and HRP2, and the antigen dynamic data from Namibia, we present a novel methodology for estimating the TTN of new RDTs. The relative contribution of HRP2 and pLDH to TTN in RDTs with combined HRP2 and LDH lines was also examined.

## Methods

### Study design and data collection

This analysis uses data from a longitudinal cohort study of 164 malaria-infected participants in the Zambezi Region of Namibia conducted in 2018, denoted herein as the primary data. In 2016, the Zambezi Region had one of the highest malaria burdens in Namibia, with an annual incidence of 30 cases per 1,000 population. The primary data are described and analyzed extensively in a paper focusing on HRP2 dynamics from Ntuku et al., 2024.^4^ In brief, the study participants consisted of a mix of symptomatic and asymptomatic individuals over 6 months of age who presented to a regional hospital, or were detected using the Standard RDT via mass screening and treatment (MSAT) campaigns, and were diagnosed with uncomplicated *P. falciparum* malaria. Individuals were excluded from the study if they displayed signs of severe malaria, had febrile conditions due to a disease other than malaria, or were otherwise ineligible to receive the standard-of-care malaria treatment artemether-lumefantrine. Enrolled individuals were treated and followed up for a maximum of 132 days. Day 0 in the study represents the date of recruitment for the study participant and the first day of sampling. For study participants this may have occurred just before treatment or within 7 days of treatment. Approximately once a week, concentrations of HRP2 and pLDH were measured, and the Standard RDT and the NxTek ultrasensitive RDT (uRDT) were conducted until an individual tested negative on either RDT for two consecutive weeks.

### Blood sampling

Approximately 0.25 mL of capillary blood was collected from each malaria-infected participant into an ethylenediaminetetraacetic acid collection tube. Directly after collection, blood samples were examined using the Standard RDTs and uRDTs (Table 1). In addition, blood samples were aliquoted in cryogenic vials and stored at -80°C for quantification of malaria antigens. Parasite density was also established via *P. falciparum* -specific quantitative PCR using the *var* gene acidic terminal sequence (varATS) method^25^ on DNA extracted from 100 μL of whole blood.

**Table 1:**
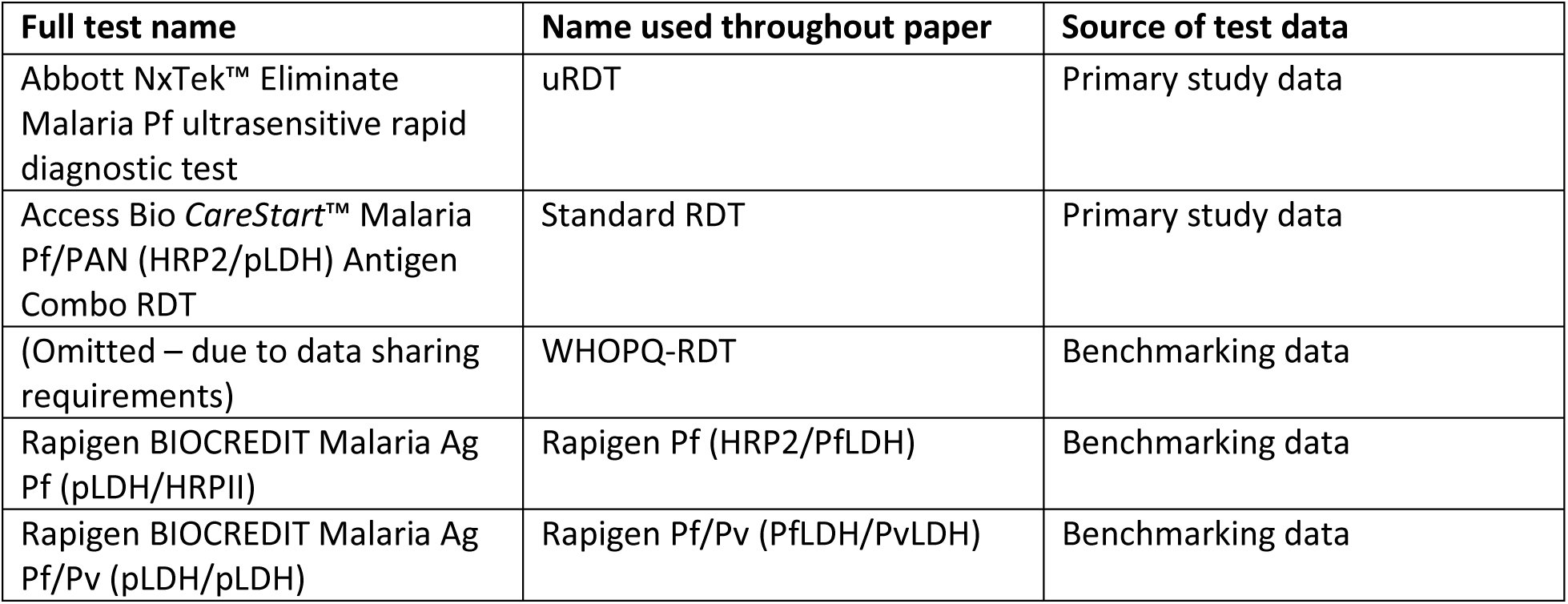
Table displaying official brand name and shortened names used throughout this paper for each RDT examined, as well as an indicator of whether the RDT results were obtained directly from primary study data or through benchmarking as described previously.^16^

| Full test name | Name used throughout paper | Source of test data |
| --- | --- | --- |
| Abbott NxTek™ Eliminate Malaria Pf ultrasensitive rapid diagnostic test | uRDT | Primary study data |
| Access Bio CareStart™ Malaria Pf/PAN (HRP2/pLDH) Antigen Combo RDT | Standard RDT | Primary study data |
| (Omitted – due to data sharing requirements) | WHOPQ-RDT | Benchmarking data |
| Rapigen BIOCREDIT Malaria Ag Pf (pLDH/HRPII) | Rapigen Pf (HRP2/PfLDH) | Benchmarking data |
| Rapigen BIOCREDIT Malaria Ag Pf/Pv (pLDH/pLDH) | Rapigen Pf/Pv (PfLDH/PvLDH) | Benchmarking data |

### Sample evaluation

Aliquots of these blood samples (n = 1,582) were shipped to the PATH laboratory (Seattle, WA) for further evaluation and stored frozen at −80°C until use. Measurement of malaria antigen concentration in 1,579 blood samples with sufficient sample volume was performed using the Quansys Biosciences QPlex™ Human Malaria (4-Plex; Quansys Bioscience, Logan, UT) which enables the user to simultaneously quantify HRP2, all-malaria lactate dehydrogenase (Pan-LDH), *Plasmodium falciparum* lactate dehydrogenase (PfLDH), *Plasmodium vivax* lactate dehydrogenase (PvLDH), and C-reactive protein. Initially, the standard testing procedure— which tests samples at neat and 1:50 dilutions—was used.^26^ Samples with high concentrations of analytes were quantified through further dilutions, if necessary.

### RDTs considered in this analysis

A summary of the RDTs included in the analysis in this article is provided in Table 1. The full proprietary test name is used to introduce each RDT on its first use in the paper, whereas the shorthand name is used in all instances thereafter.

### Benchmarking data (secondary data)

To investigate the potential TTN of next-generation RDTs, in which this type of longitudinal study has not yet been conducted, a second dataset of laboratory-based RDT benchmarking test data was used. Two RDTs were examined in the benchmarking: the Rapigen BIOCREDIT Malaria Ag Pf/Pv (pLDH/pLDH) test, containing test lines to detect PfLDH and PvLDH, and the Rapigen BIOCREDIT Malaria Ag Pf (pLDH/HRP II) test, with distinct PfLDH and HRP2 detection lines, herein referred to as the Rapigen tests (Table 1). Panels used for testing were prepared by dilution of recombinant proteins, cultured malaria parasites, international standards, or clinical samples into malaria-negative donor blood, which were then aliquoted and frozen. All panel members were tested using the Q-Plex array to quantify malaria antigen concentrations.^16^ Tests were run according to instructions for use with venous whole blood. Each panel member was tested with three to five replicate RDTs for all panel proteins used until a clear pattern of negativity was reached with lower concentrations of two or more adjacent dilutions testing negative for 100 percent of replicates. Concentrations near the LOD were chosen for specific panel members and run with a total of 40 replicate tests that were above, at, and below the concentration identified as near the LOD—except for panel members containing human recombinant lactate dehydrogenase (LDH), clinical pool dilutions, clinical individual samples, or culture-derived panel members with limited quantities. Test-line intensity was assigned based on a comparison to an intensity scale card provided by the manufacturer. Any visible test line was considered positive. All test results were interpreted according to manufacturer instructions.

Logistic regression models were then fitted with RDT positivity as the dependent variable and (log10-transformed) antigen concentration as the independent variable. From these model fits, the antigen concentration at which there is a 90 percent probability of positivity was determined, and this was deemed the threshold detectable concentration. For RDTs that only used a single antigen for a given *Plasmodium* species, a regression model was used. For RDTs that had two analytes for *P. falciparum* (i.e., HRP2/PfLDH tests), two separate logistic regression models were run, and the outputs were combined on a surface to give an estimated probability that either (or both) test lines would be positive based on both HRP2 and PfLDH antigen concentrations. Statistical analyses were conducted using R version 4.2.1 software.^27^ Further model details are presented in Golden et al., 2023.^16^

### Antigen decay models

Linear mixed-effect models were fitted to the Namibia data to estimate both the monophasic and biphasic decay rates of pLDH posttreatment using criteria that minimized both Akaike information criterion (AIC) and Bayesian information criterion (BIC). The monophasic decay model assumed a constant rate of decay over time, whereas the biphasic model allowed for two separate rates of decay before and after a designated switch point, with the initial decay presenting as short and rapid, followed by a longer period of slower decay.

The functional forms of the monophasic and biphasic exponential decay models are as follows: Monophasic decay:

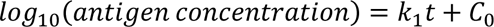

Biphasic decay:

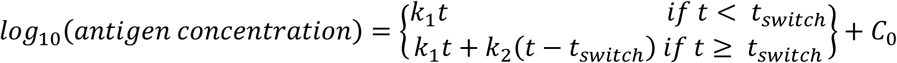

where t = time posttreatment (days), k_1_ and k_2_ are the distinct decay parameters before and after t_switch_ (the switch point), and C_0_ is the initial log10-transformed antigen concentration.^24^ Individual-level random effects were included in the models to account for differing levels of initial antigen concentration for each participant in the study. The switch point was determined by iterating the biphasic models over a range of possible times and selecting the one that minimized the AIC and BIC.

### Time to negativity models for next-generation RDTs

Kaplan-Meier (KM) survival curves were used to estimate the TTN of next-generation RDTs in three different ways. Firstly, we wanted to understand the TTN of next-generation RDTs with different hypothetical LODs of 50, 100, and 1,000 (and 5,000 and 10,000 for pLDH only) pg/mL. To do this we used the longitudinal antigen-concentration data from the Namibia dataset and calculated the proportion of individuals that would have antigen levels lower than the LOD thresholds at each time point and therefore be RDT negative. These data were then converted into a KM survival curve to estimate TTN for different RDT LODs.

Secondly, we wanted to estimate the TTN of the new Rapigen RDTs and compare them to TTN for the uRDT and the Standard RDT used in the Namibia study. For the Standard RDT and uRDT, KM curves were simply derived from the data directly. For the Rapigen RDTs, we used LOD estimates generated in the RDT benchmarking,^16^ and similar to the hypothetical LOD approach, calculated the proportion of the participants from the Namibia data that had antigen concentration levels below these LODs. In the case of the Rapigen Pf (HRP2/PfLDH) RDT with both HRP2 and PfLDH lines, an individual was considered negative only when both HRP2 and PfLDH concentrations fell below their respective LODs. These data were then converted into KM survival curves and compared to the RDTs used in the study.

Finally, we wanted to understand the relative contribution of HRP2 and PfLDH in the TTN for a widely used WHO prequalified RDT (WHOPQ-RDT, HRP2/PfLDH) and the Rapigen Pf (HRP2/PfLDH) RDT. To do this, for each RDT, we calculated the proportion of participants in the Namibia study that had antigen concentrations below the LOD for each antigen separately and then below the LOD for both antigens. These data were then used to generate KM curves for the two RDTs based on each antigen alone and both antigens in concert.

## Results

### Study population

The study population consisted of 164 Standard RDT–confirmed *P. falciparum* -positive patients from the Zambezi Region in the northeast of Namibia. Of these individuals, 29 were excluded from the TTN and antigen decay models described above: 15 were excluded from this analysis due to posttreatment resurgence in pLDH that may have indicated possible reinfection or treatment failure, and 14 were excluded because they had no polymerase chain reaction data collected to confirm parasite presence. Additionally, 2 individuals were excluded from the original study population of 164 because they left the study while still positive for malaria by Standard RDT. The full flowchart of participant exclusions is shown in Supplementary Figure 1.

Of the 135 individuals included in the analysis, the mean age was 20.7 years (standard deviation of 14.5 years), with ages ranging from less than 1 year to 80 years. A total of 11 individuals (8.1 percent) were under the age of 5 years, 62 individuals (45.9 percent) were female, and 19 individuals (14.1 percent) were recruited to the study via MSAT campaign.

Profiles of the parasite density, HRP2 and pLDH concentration for all study participants are provided in Supplementary Figure 2 along with the age and gender listed for each participant. In 10 of the 15 cases where the pLDH concentration rose again after the initial drop posttreatment, a persistence or increase in parasite density was observed by quantitative PCR in the samples.

For the 62 individuals for which sample was collected prior to treatment the geometric mean antigen concentrations for HRP2 and pLDH were 899 ng/mL (95% CI: 544 - 1254) and 344 ng/mL (95% CI: 342 - 347).

### Antigen decay model results

Monophasic and biphasic exponential decay models were fitted to the HRP2 and pLDH antigen data for all participants in the final analysis subset. A biphasic exponential decay model provided the best estimate of antigen dynamics over time for both HRP2 and pLDH, outperforming monophasic models as evidenced by lower AIC and BIC values that are displayed in Supplementary Table 1. The final fits for both antigens are shown in Figure 1.

**Fig. 1.**
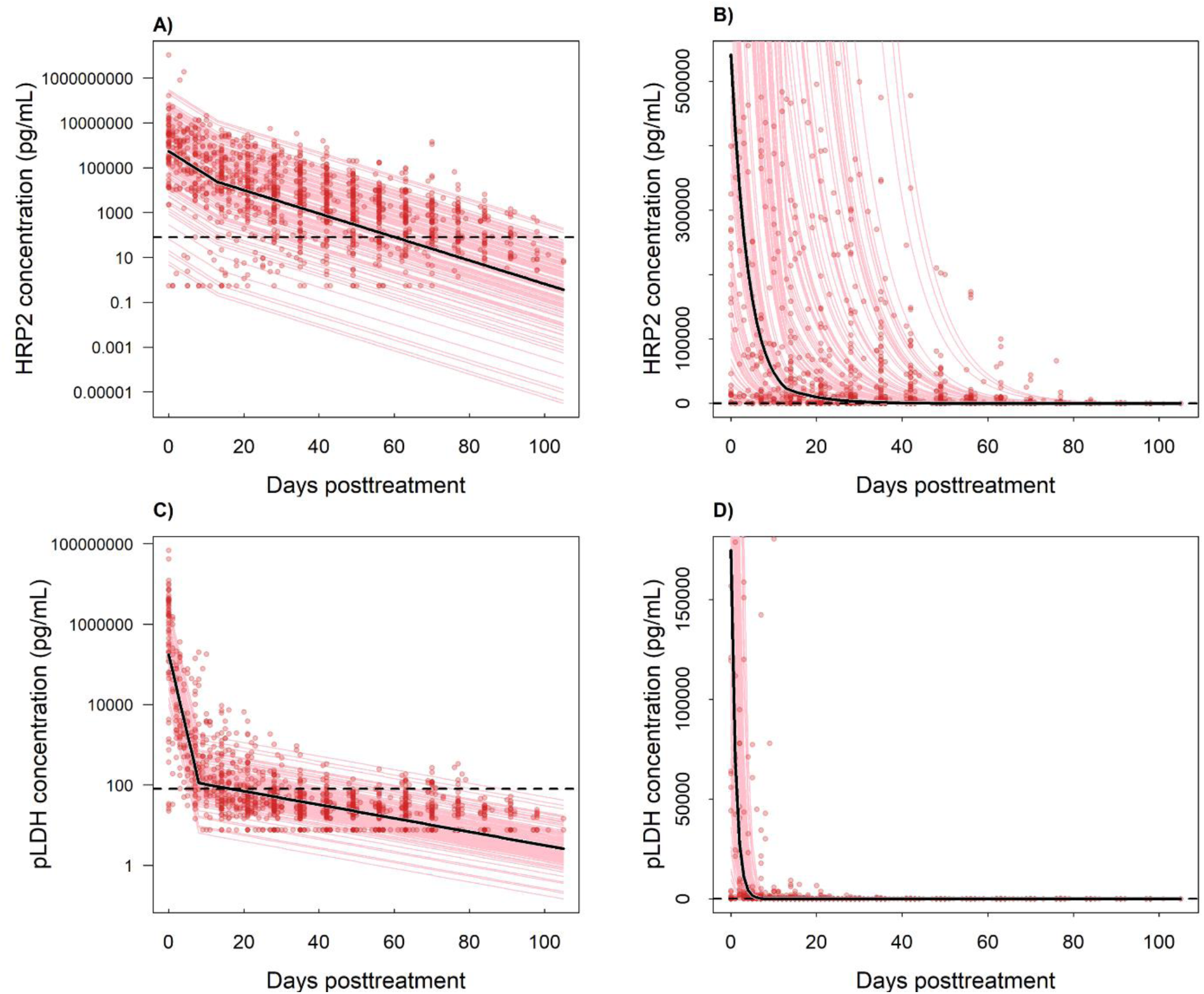
HRP2 and pLDH antigen dynamics posttreatment. HRP2 data reported first in Ntuku et al., 2024.^4^ Red points indicate the data from each individual over time, the pink lines show the fitted biphasic decay for each participant, and the solid black lines show the overall fit to all the data. Dashed black lines indicate the uRDT LOD value of 80 pg/mL. Panels A and C present the decay on a log10 scale, and panels B and D show the untransformed data. The y-axis for panels B and D is restricted to amplify the biphasic inflection.

The decay parameters were k1 = –0.105 and k2 = 0.053, and the switch point (*t*_*switch*_) was day 13 for the HRP2 model. For the pLDH model, the final parameters were k1 = –0.399 and k2 = 0.382 with a switch point *t*_*switch*_ of 8 days. These model fits show there is a substantially more rapid decay in pLDH than in HRP2, as shown previously.^24^ Model predictions showed that HRP2 decayed to half its initial concentration after 3.9 days, whereas pLDH only took 1.8 days to reach half its initial concentration. Furthermore, the model indicates that it takes an average of 61 days to reach levels of < 80 pg/mL for HRP2, which equates to the approximate LOD of the uRDT.^11^ This is compared to an average of 16 days for pLDH to reach < 80 pg/mL in this study population. These data are shown in the histograms of the time posttreatment required to reach 80 pg/mL in Supplementary Figure 3. The equivalent times to reach undetectable levels for current RDTs, assuming estimated LODs of < 1,000 pg/mL for HRP2 and < 5,000 pg/mL for pLDH, were 44 days and 4 days, respectively.

### RDT time to negativity

The first TTN analysis shows the KM survival curves assuming different hypothetical LODs of 50, 100, or 1,000 pg/mL of both antigens, as well as 5,000 and 10,000 pg/mL for pLDH (Figure 2). For HRP2, with a LOD of 1,000 pg/mL, the estimated median TTN was 49 days. If improved-sensitivity RDTs with LODs of 100 or 50 pg/mL were available, we estimate the median TTN would increase by 14 days. For pLDH, at high LODs, the median TTN is 7 days. A new RDT with a LOD of 1,000 pg/mL (similar to the estimated LOD of the new Rapigen RDTs) would increase the median TTN to 9 days. Further pLDH LOD improvements to 100 or 50 pg/mL are estimated to increase the median TTN to 18 and 21 days, respectively.

**Fig. 2.**
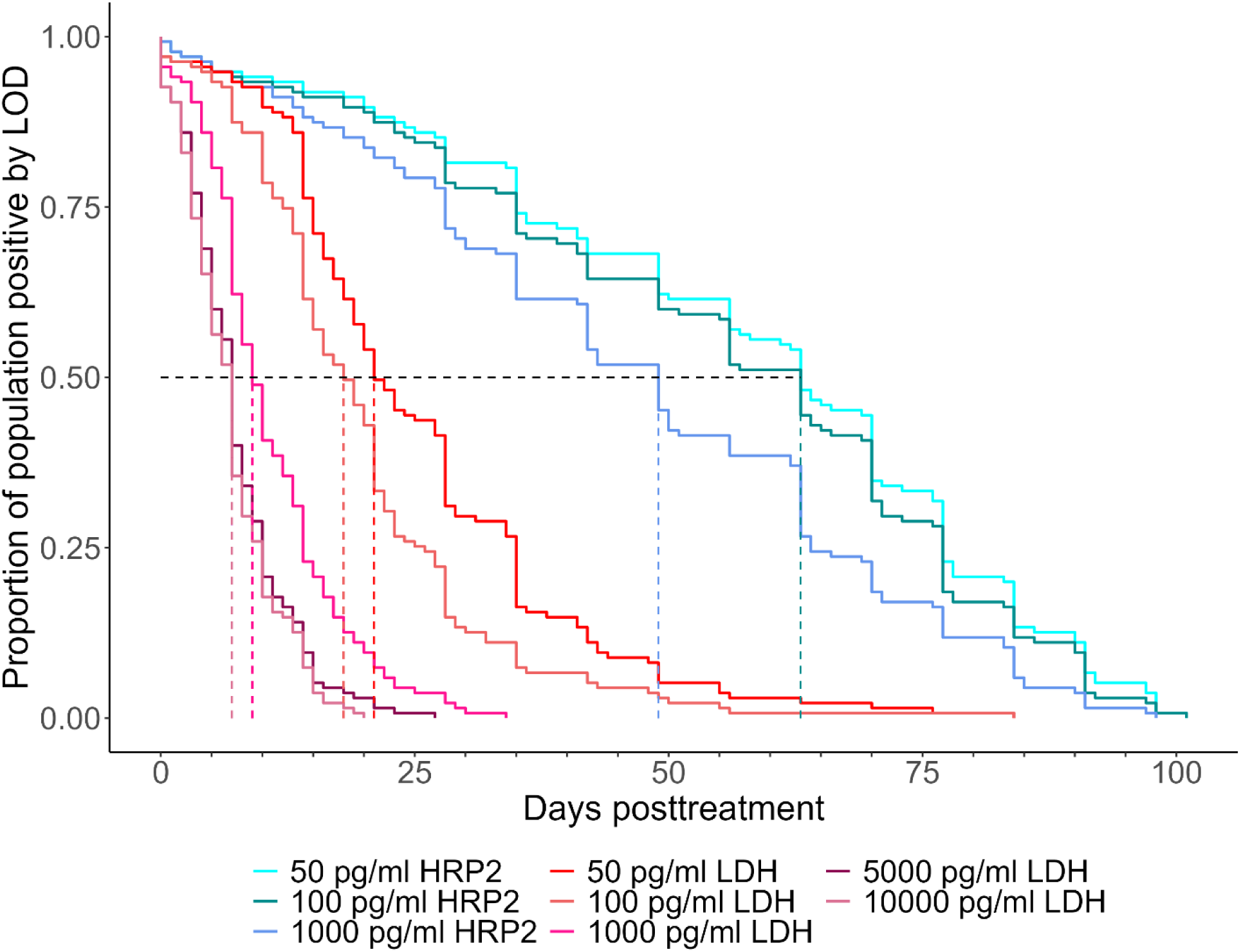
Combined survival curve for different hypothetical limits of detection (pg/mL) for each antigen. pLDH measurement lines are in shades of red, and HRP2 measurement lines are in shades of blue. Dashed lines represent the median TTN for each test line.

Figure 3 shows the observed KM survival curves for the uRDT and the Standard RDT used in the Namibia study, as adapted from results first reported in Ntuku et al., 2024.^4^ The median TTN for the uRDT was estimated at 68 days posttreatment compared to 42 days for the Standard RDT. These are compared against KM survival curves for a different, but widely used WHO prequalified RDT, referred to here as WHOPQ-RDT—and the Rapigen RDTs. To generate the comparison, the LODs as quantified elsewhere^16^ and listed in Supplementary Table 2 are used to calculate the proportion of individuals in the Namibia dataset with antigen concentrations below these LODs, and thus presumably RDT negative, each day posttreatment. The resulting estimated median TTNs were 49 days for the WHOPQ-RDT, 56 days for the Rapigen Pf (HRP2/PfLDH) RDT, and 10 days for the pLDH-based Rapigen Pf/Pv (PfLDH/PvLDH) RDT (Figure 3).

**Fig. 3.**
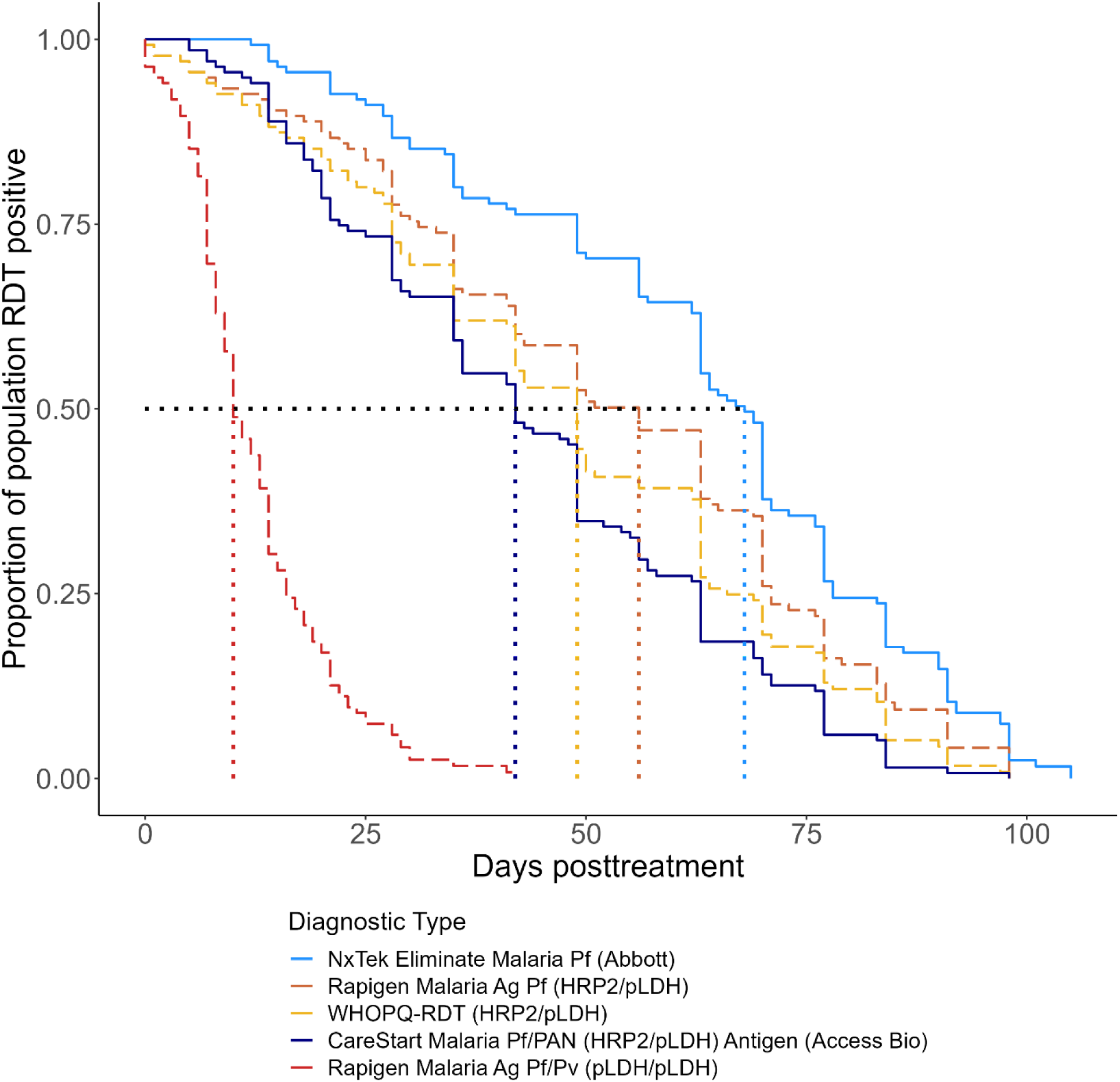
Survival curves for NxTek, *CareStart*, WHOPQ-RDT, and Rapigen Malaria Ag RDTs among participants who cleared infection. Dotted lines represent the median TTN for each RDT. The solid blue NxTek and *CareStart* lines were calculated using primary data from this study reported first in Ntuku et al., 2024.^4^ The dashed red, orange, and yellow lines were estimated using RDT benchmarking data published in Golden et al., 2023.^16^

Finally, we estimated the TTN based on only the HRP2 test line or only the pLDH test line of the WHOPQ-RDT and Rapigen Malaria Ag RDTs. The results show that the HRP2 components are predicted to remain positive for 49 and 57 days, respectively, whereas the pLDH components are predicted to remain positive for 7 and 9 days, respectively (Figure 4).

**Fig. 4.**
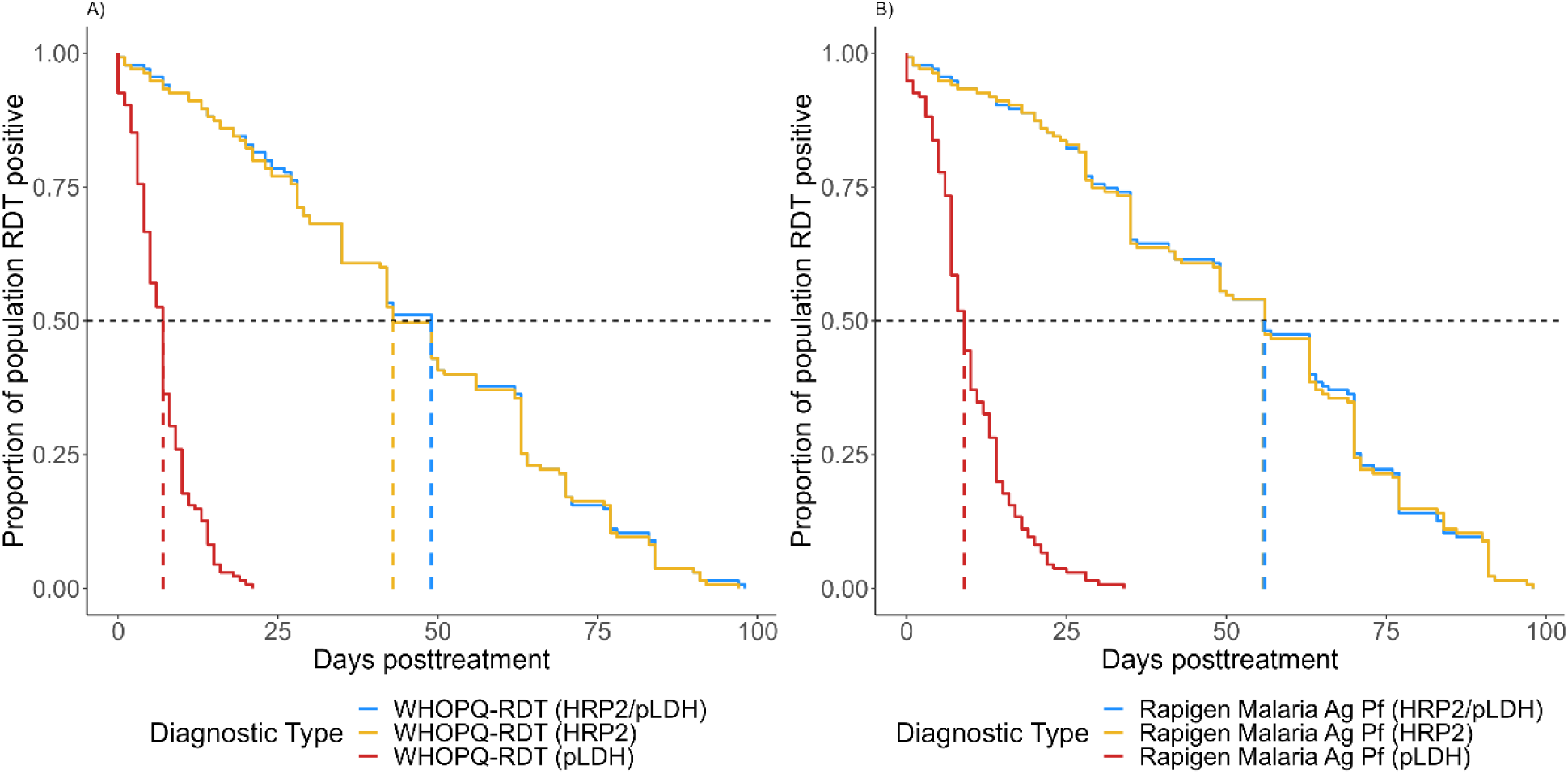
Survival curves for WHOPQ-RDT and Rapigen Malaria Ag Pf RDT among participants who cleared infection. The WHOPQ-RDT is shown in panel A, while the Rapigen Malaria Ag Pf RDT is shown in panel B. Blue lines indicate the combined HRP2/PfLDH trend, while yellow and red lines represent the HRP2 and PfLDH-only lines, respectively. Dashed lines represent the median TTN for each RDT/test line.

## Discussion

The TTN for RDTs informs our understanding of how RDTs ensure appropriate clinical case management, especially for individuals presenting with symptoms a short time after being treated for a prior infection. The TTN is also important in understanding the results from RDT-positivity rates in cross-sectional surveys and interventions, such as MSAT. In this analysis, we present a novel quantitative framework for assessing the TTN of new RDTs using previously collected longitudinal data. Underpinning this framework is: (i) an accurate measure of the analytical sensitivity of a given RDT for the antigen(s) it is measuring in terms of antigen concentration as determined by standardized methods,^16^ and (ii) characterization of longitudinal specimens characterized for antigen concentration.

Firstly, we fitted biphasic decay models to capture the dynamics of HRP2 and pLDH antigens over time. This confirmed previous findings that pLDH has a shorter half-life than HRP2.^22,28,29^ We then estimated the median TTN for different hypothetical LODs for both HRP2 and pLDH. This showed that HRP2-based RDTs have a longer TTN than LDH-based RDTs, and new RDTs with lower LODs, such as the uRDT from this study, increase this duration by an additional 14 days. However, for pLDH-based RDTs, current models have a very short median TTN, estimated at 7 days, and new tests with LODs of around 1,000 pg/mL are estimated to increase the TTN to 9 days. Applying LODs quantified in a previous benchmarking study for two new Rapigen Malaria Ag RDTs^16^ estimated that the test with both HRP2 and pLDH lines would have a median TTN of 56 days, whereas the Rapigen Pf/Pv (PfLDH/PvLDH) RDT would have a median TTN of 10 days for *P. falciparum*.

This analysis also provided a means to validate our TTN estimation approach. Figure 3 compares the Standard RDT conducted on posttreatment patients in Namibia against estimates derived by combining the Namibia antigen data with laboratory-generated LOD estimates for a different prequalified test the WHOPQ-RDT. Here we see the TTN estimates are comparable by the two approaches (42 versus 49 days). Similarly, the TTN of the uRDT used in the Namibia study can be compared to the TTN estimates generated for the Rapigen Pf (HRP2/PfLDH) RDT. These values are 68 and 56 days, respectively, and consistent with known HRP2 LODs of 80 pg/ml for the uRDT compared to ∼525 pg/mL for the Rapigen Pf test.^30^

Conducting longitudinal studies for all new RDTs being developed is time consuming and expensive. Quantifying the TTN is not a requirement for WHO prequalification of a new RDT, so manufacturers typically do not conduct them, leaving the work to researchers^31^ and national malaria programs.^32^ Therefore, leveraging longitudinal cohort data with our method can inform how the LOD of a new test corresponds to its TTN.

With the emergence of *hrp2/hrp3* deletions,^33^ there has been a renewed interest in RDTs that target the pLDH antigen. Until recently, there was an almost exclusive preference for HRP2-based RDTs for *P. falciparum* due to their better performance in terms of sensitivity in populations where there is no *hrp2/hrp3* deletion, as well as often consistently improved shelf-life stability profiles. However, in regions where these deletions are present, national malaria control programs have started to shift procurement to pLDH-based RDTs^17,34^—and with this comes a need for higher-sensitivity pLDH-based RDTs to provide similar diagnostic performance to the HRP2-based RDTs. Two products with significantly improved analytical sensitivity for pLDH are the Rapigen Pf (HRP2/PfLDH) and the Rapigen Pf/Pv (PfLDH/PvLDH).^16^ The Rapigen Pf/Pv (PfLDH/PvLDH) has been shown to perform better in populations with *hrp2/hrp3* deletions compared to RDTs that have HRP2 and pLDH (with lower sensitivity).^21^ The Rapigen Pf/Pv (PfLDH/PvLDH) is already being used programmatically or considered for such, in Ethiopia, Eritrea and Djibouti to minimize false negative RDTs arising from *hrp2/hrp3* deletions.

The study also analyzed the performance of tests which detect both HRP2 and PfLDH. While currently there are only RDTs with separate lines for both antigens, RDTs with both HRP2 and LDH detection on the same line are anticipated in the near future. These combined line tests have the operational benefit over separate line tests in that they look like and are interpreted by the end user in exactly the same way as the currently widely used HRP2 line only RDTs for diagnosing *P.falciparum* infection regardless of an underlying hrp2/hrp3 deletion. The results presented here show that for tests with dual lines for HRP2 and PfLDH, the overall TTN of the test is strongly driven by the HRP2 line. This is driven by both the higher starting concentration of HRP2 and the slower decay rate of the HRP2 antigen post-treatment compared to those of PfLDH. In *P. falciparum* RDTs where the PfLDH line and the HRP2 lines are separate we can anticipate that the PfLDH line will be more indicative of an active or very recent infection (within the past ten days) versus the HRP2 line which persists for much longer. In an RDT with combined HRP2/PfLDH lines, positivity will most likely reflect that of the persistent HRP2 antigen. In a study in Burundi, the PfLDH line in the Rapigen Pf (HRP2/PfLDH) did also contribute to the overall incremental sensitivity of the test compared to an HRP2-only test, as did the HRP2 line on the Rapigen test which was also more sensitive (but less specific) than the HRP2 line for the comparator.^15^ The Burundi samples were active infections where higher relative concentrations of PfLDH are expected.^19^

This analysis assumes the antigen decay dynamics of the Namibian study participants are representative of symptomatic malaria patients more broadly, but there are several reasons why this might not be the case. Namibia is a country with very low malaria transmission, meaning that malaria-infected individuals are likely to have lower pre-existing immunity, and therefore may mount different immune responses upon infection compared to an individual that resides in a high-transmission area. However, the overall relative decay rates for HRP2 and LDH presented here are similar to those found previously in a setting with significantly higher transmission rates in Mali.^24^ This study also assumes that the LODs for new RDTs generated in the benchmarking process are reflective of LODs that would be observed in a field setting. This hypothesis can only be tested by generating LODs for RDT using both methods; however, the similarity in the TTN of two standard RDTs tested by two different methods (Figure 3) gives us confidence that our quantitative approach is providing valid estimates.

Several studies have shown persistence of positivity in HRP2-based RDTs more than 28 days posttreatment.^24,29,35,36,37^ However, a 2018 literature review conducted by Dalrymple et al.^22^ showed that 50 percent of patients present a negative test by day 19 (CI: 10–31) when an artemisinin-based combination therapy is given on day 0, making the TTNs for HRP2-based tests in this study more durable than have generally been observed. The TTNs for pLDH-based tests calculated in this study are generally greater than those seen in the literature which are often closer to 2 days than the 7 observed here.^22,29^

Overall, this work provides the first estimates of the TTN of two new malaria RDTs and a methodology for estimating the TTN for future malaria RDTs. By exploring the relationship between antigen dynamics, RDT LODs, and the TTN, we have provided evidence to inform the target product profile of new malaria RDTs, highlighting that even considerably more sensitive pLDH-based RDTs are unlikely to have the same long TTNs of current HRP2-based RDTs based on the decay rates of the two antigens. As evidence is generated indicating the spread of *hrp2/hrp3*-deleted parasites across malaria-endemic regions, understanding the performance characteristics of pLDH-based RDTs, including those that combine HRP2 and pLDH on the same test either through a common line or on separate lines, will become more important in order to support national malaria control program decision-making around RDT use.

## Data Availability

After publication, data collected from this study are available upon request to the corresponding author. Available data include de-identified individual participant data and a data dictionary defining each field in the set. Requests to conduct analyses outside the scope of this publication will be reviewed by the principal investigators (MSH and DM) to determine whether a proposed use of the data is scientifically and ethically appropriate and does not conflict with constraints or informed consent limitations identified by the institutions that granted ethical approval for the study. Requests to reanalyse the data presented in this Article will not require such review.

## Abbreviations

AIC: Akaike information criterion
BIC: Bayesian information criterion
HRP2: histidine-rich protein 2
KM: Kaplan-Meier
LDH: lactate dehydrogenase
LOD: limit of detection
MOHSS: Ministry of Health and Social Services
MSAT: mass screening and treatment
Pan-LDH: all-malaria lactate dehydrogenase
Pf: *Plasmodium falciparum*
PfLDH: *Plasmodium falciparum* lactate dehydrogenase
pLDH: *Plasmodium* lactate dehydrogenase
PvLDH: *Plasmodium vivax* lactate dehydrogenase
RDT: rapid diagnostic test
TTN: time to negativity [in the title]
uRDT: ultrasensitive rapid diagnostic test
WHO: World Health Organization

## Supplementary Materials

**Supplementary Figure 1:**
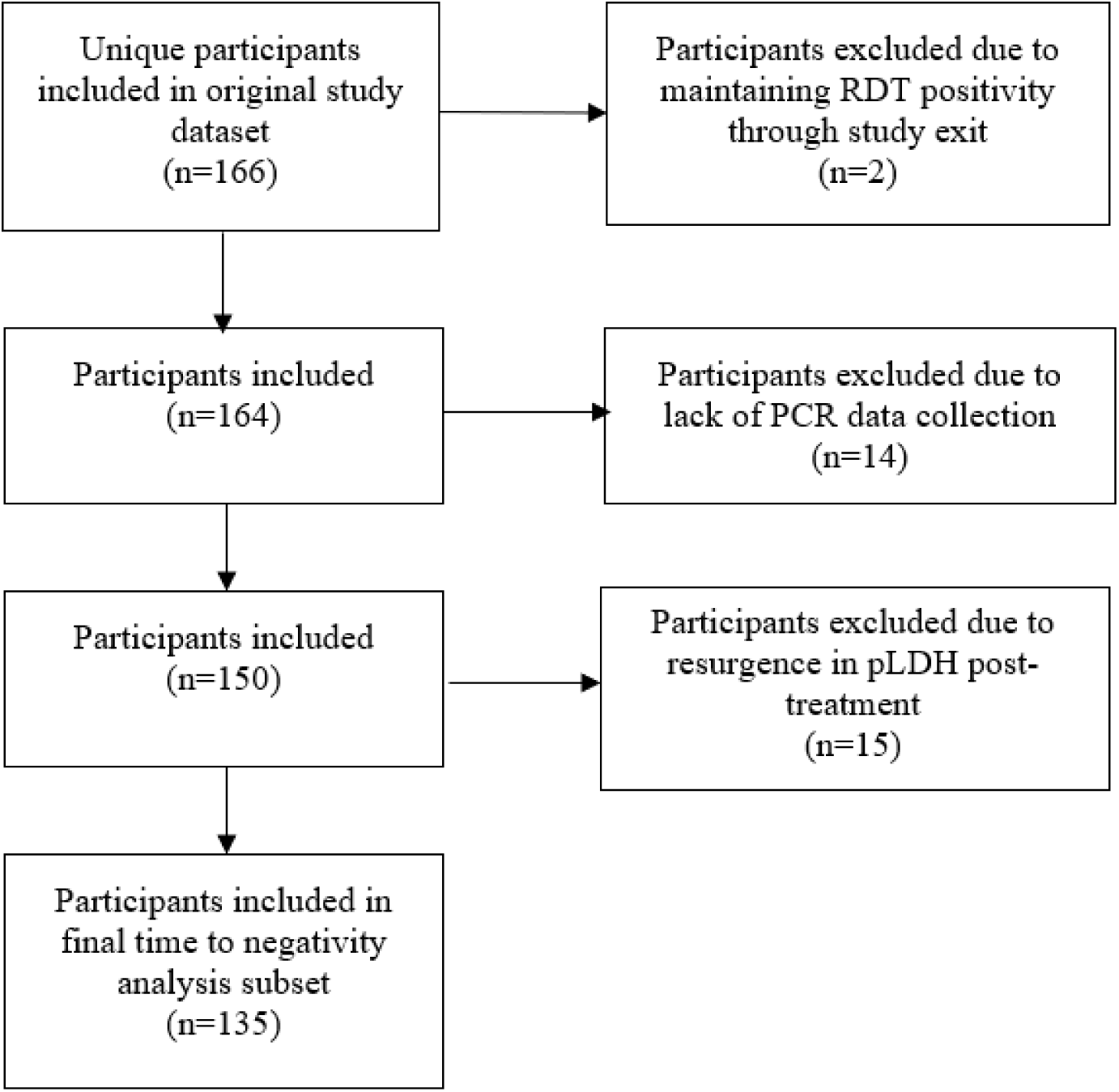
PRISMA-flow style diagram depicting exclusion and inclusion decision flow for participants from the initial study by Ntuku et al, 2024.^4^ Numbers differ from final inclusion in Ntuku et al due to separate requirements for individuals to have begun submitting samples on Day 1 of enrollment in study.

**Supplementary Figure 2:**
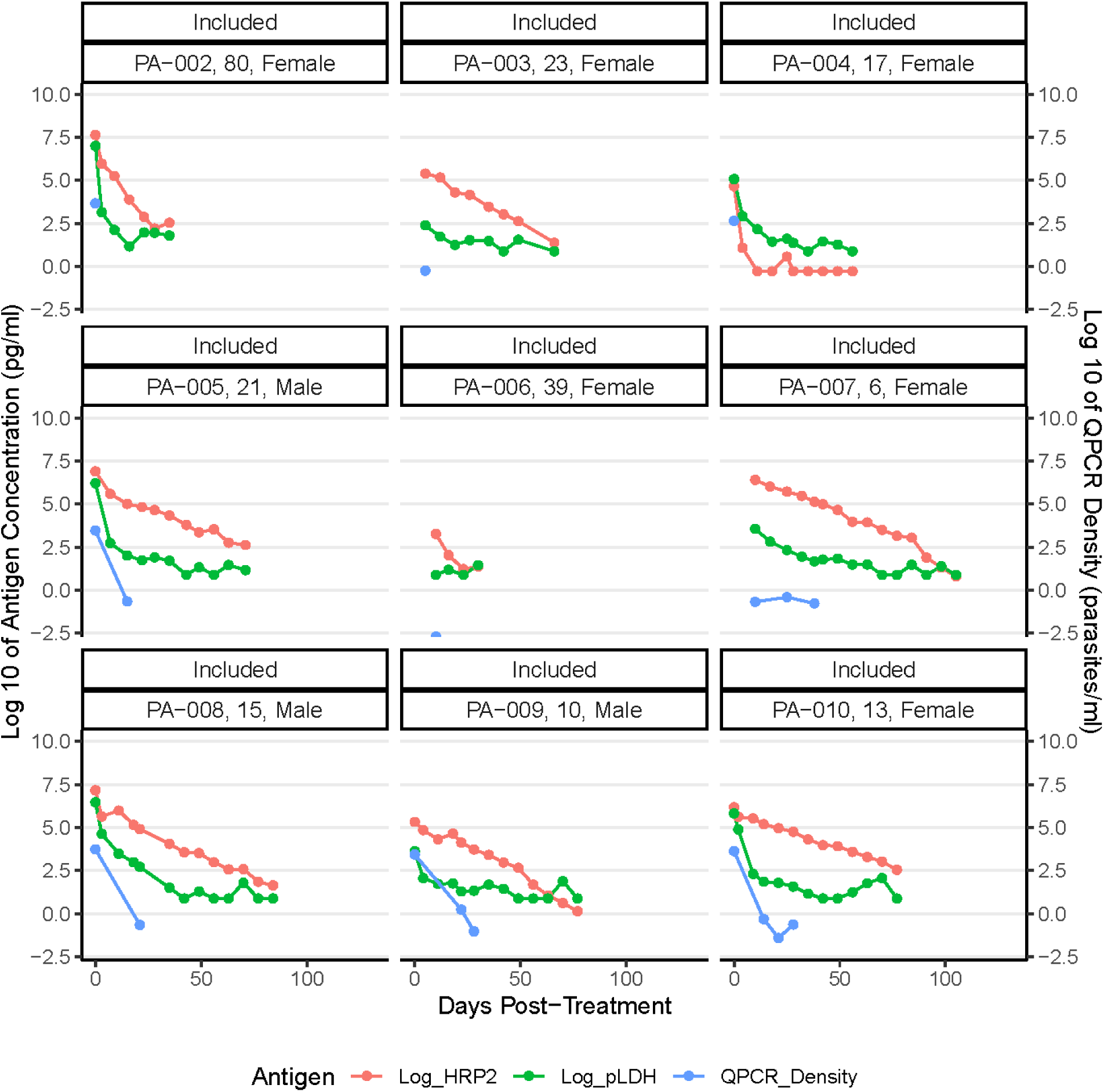
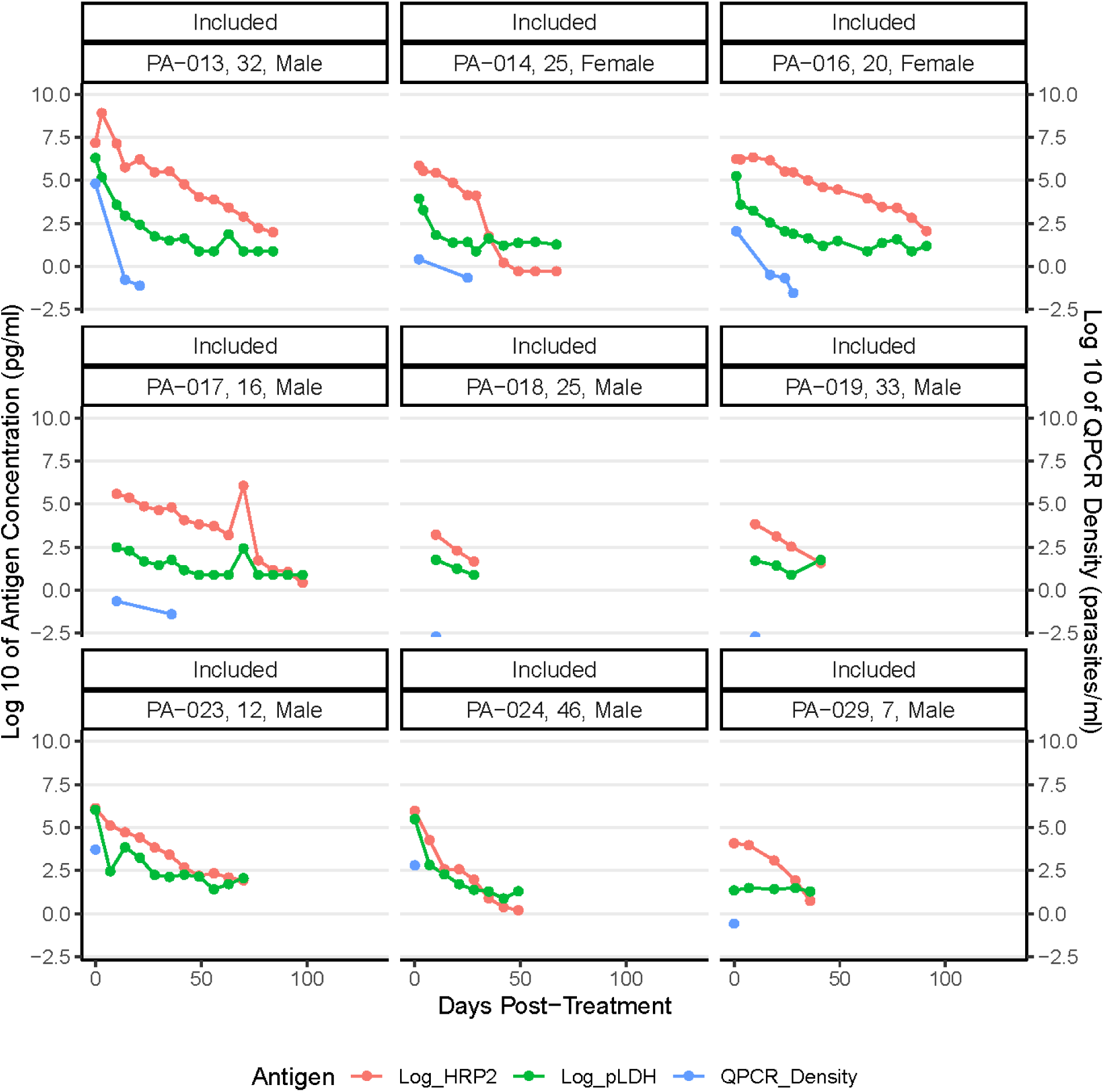
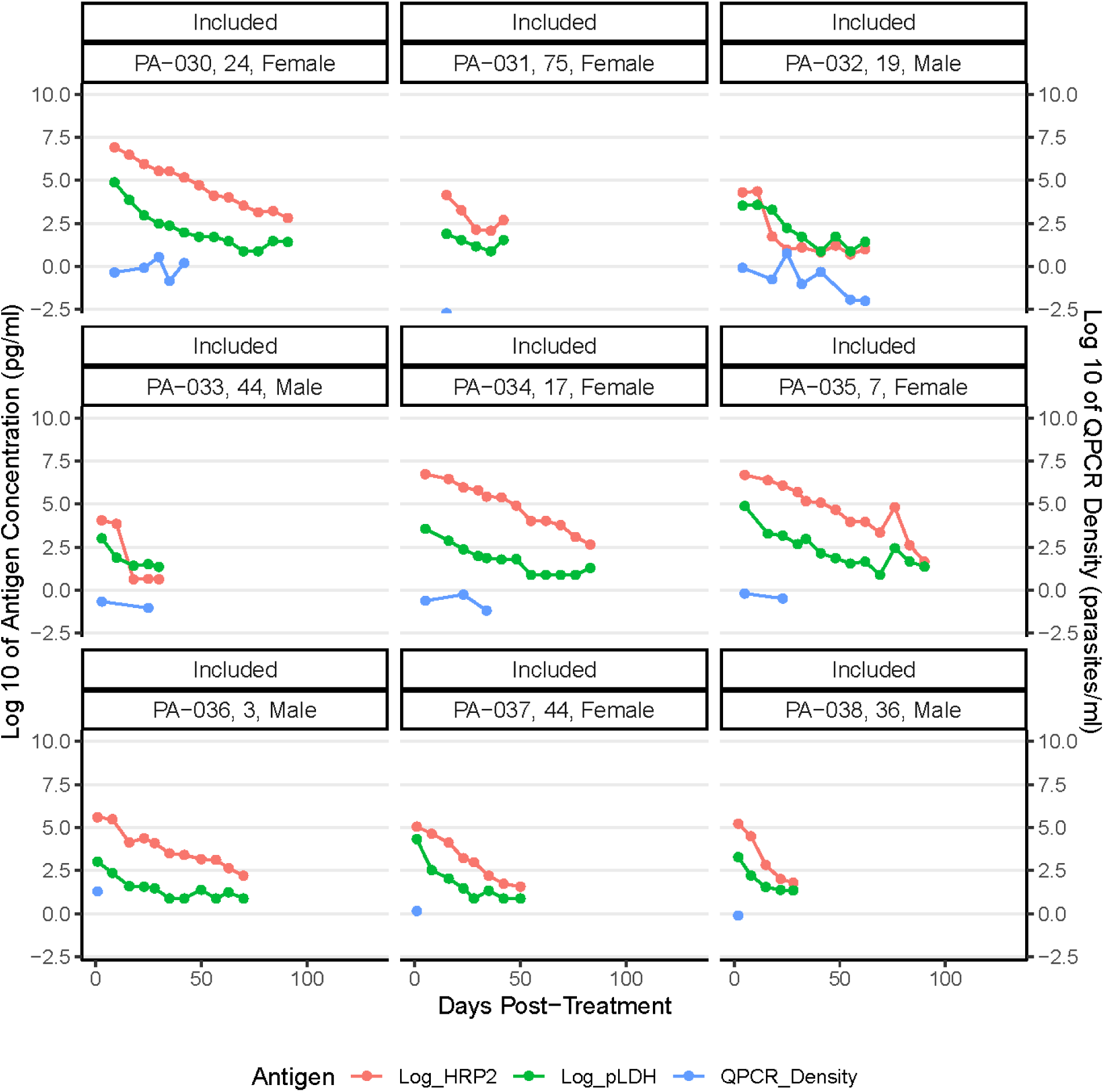
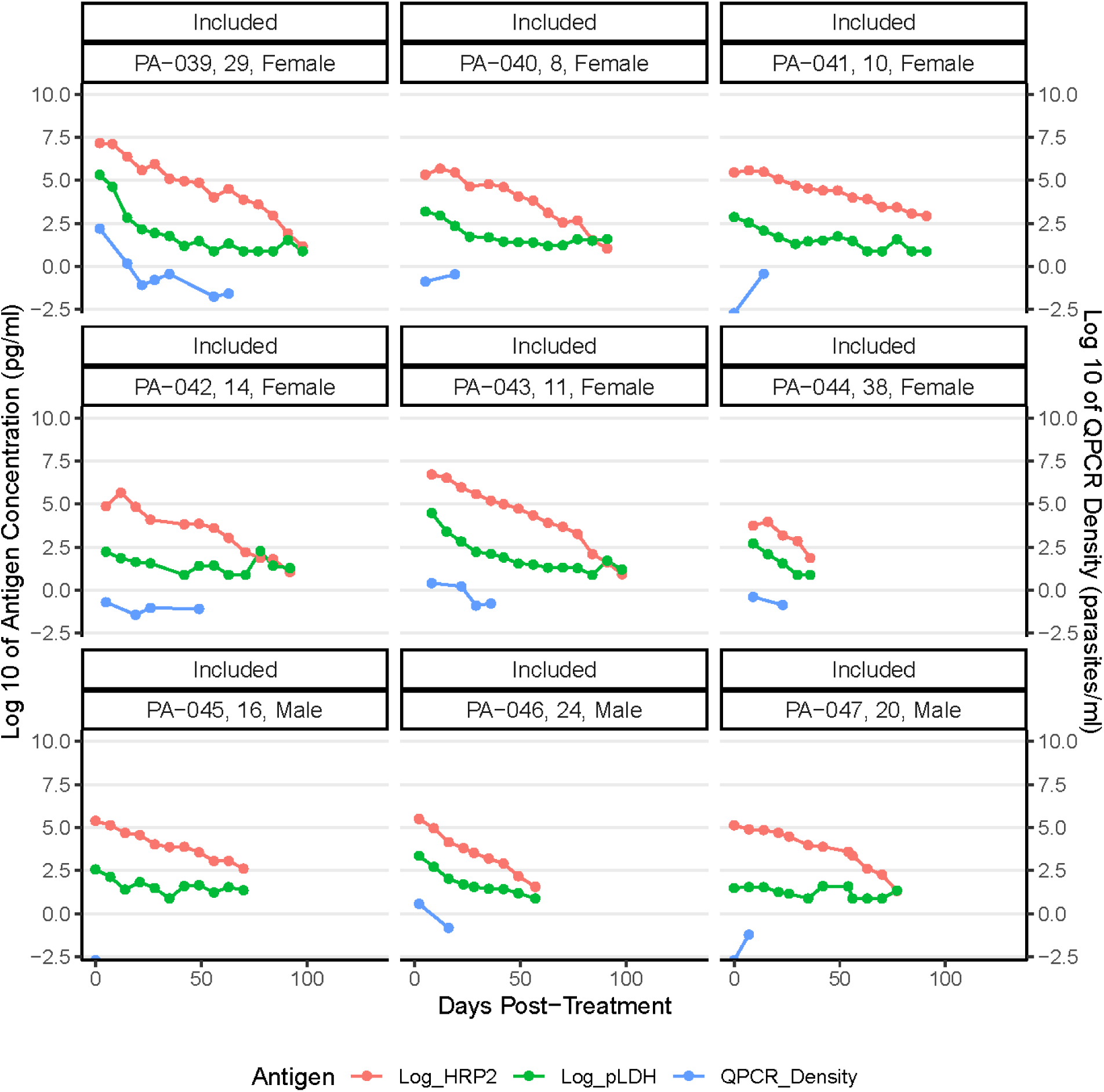
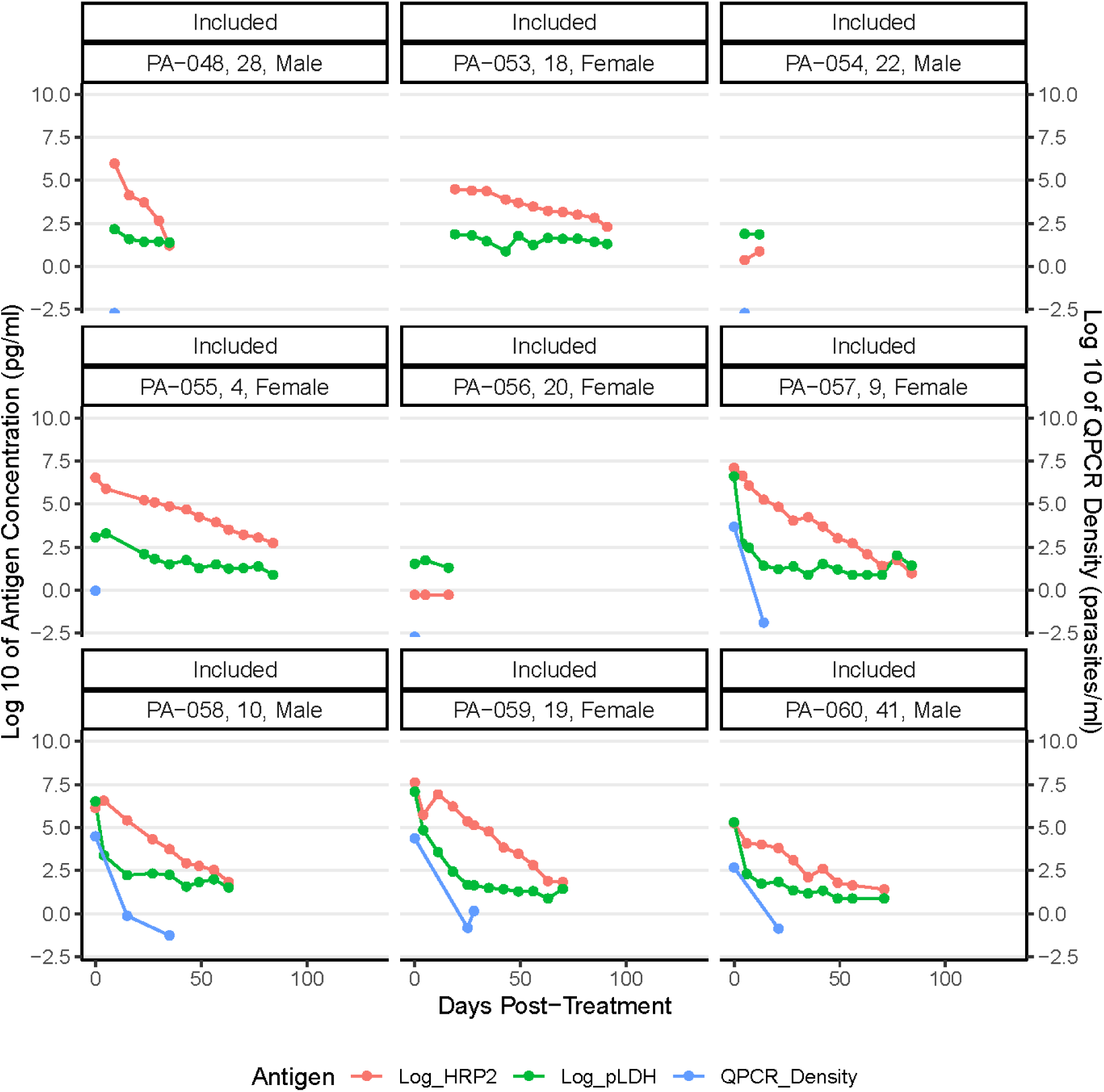
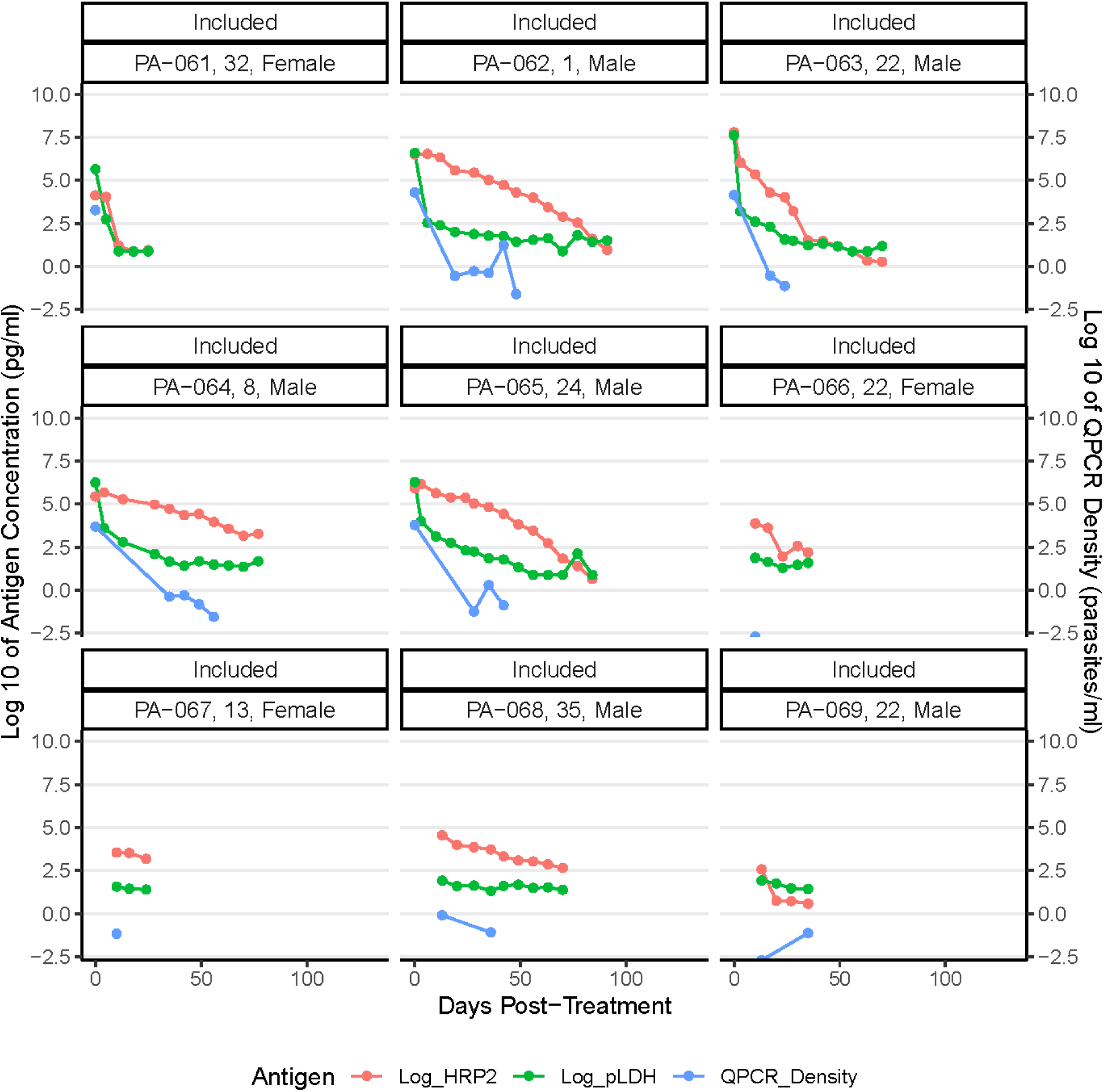
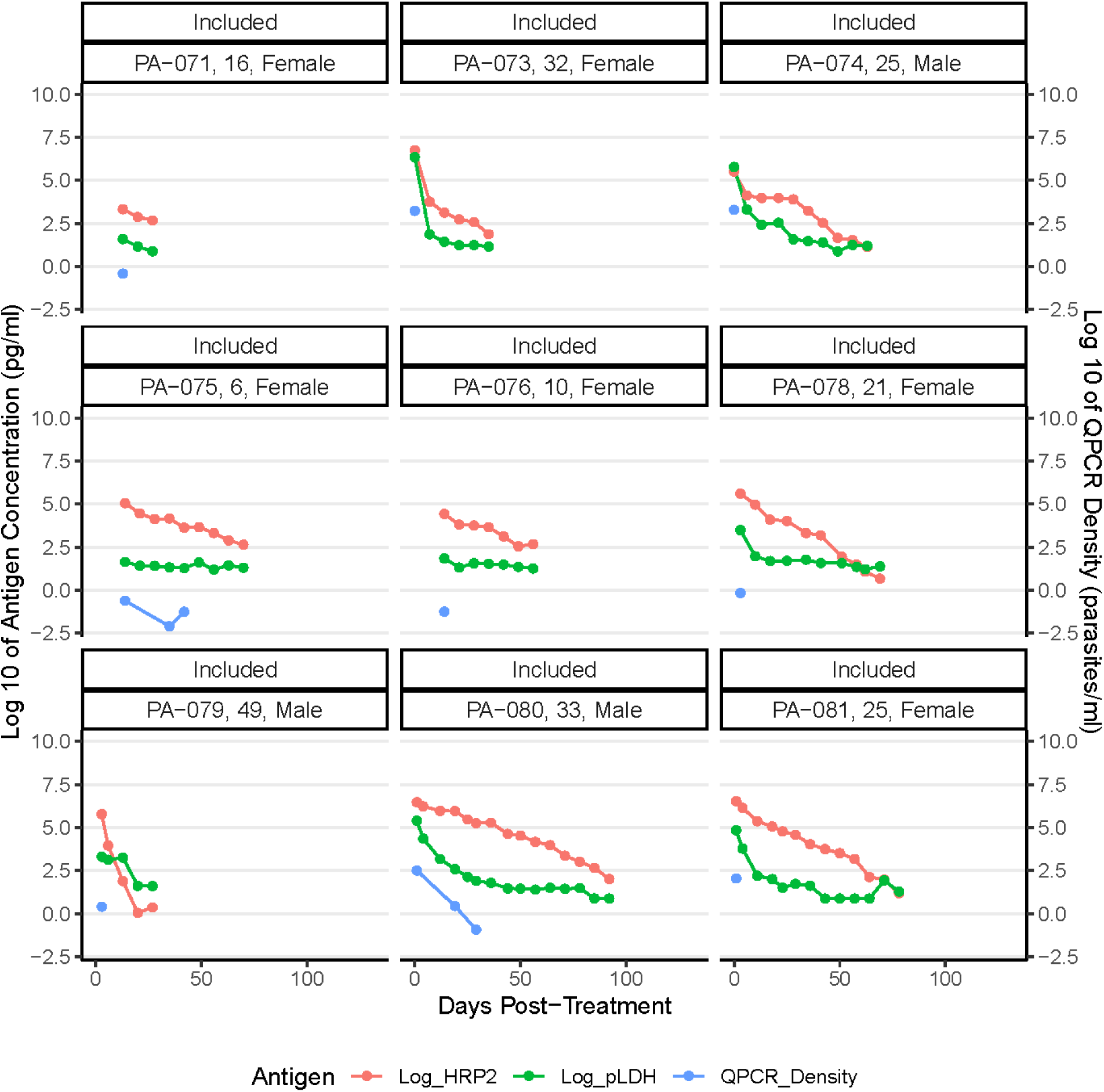
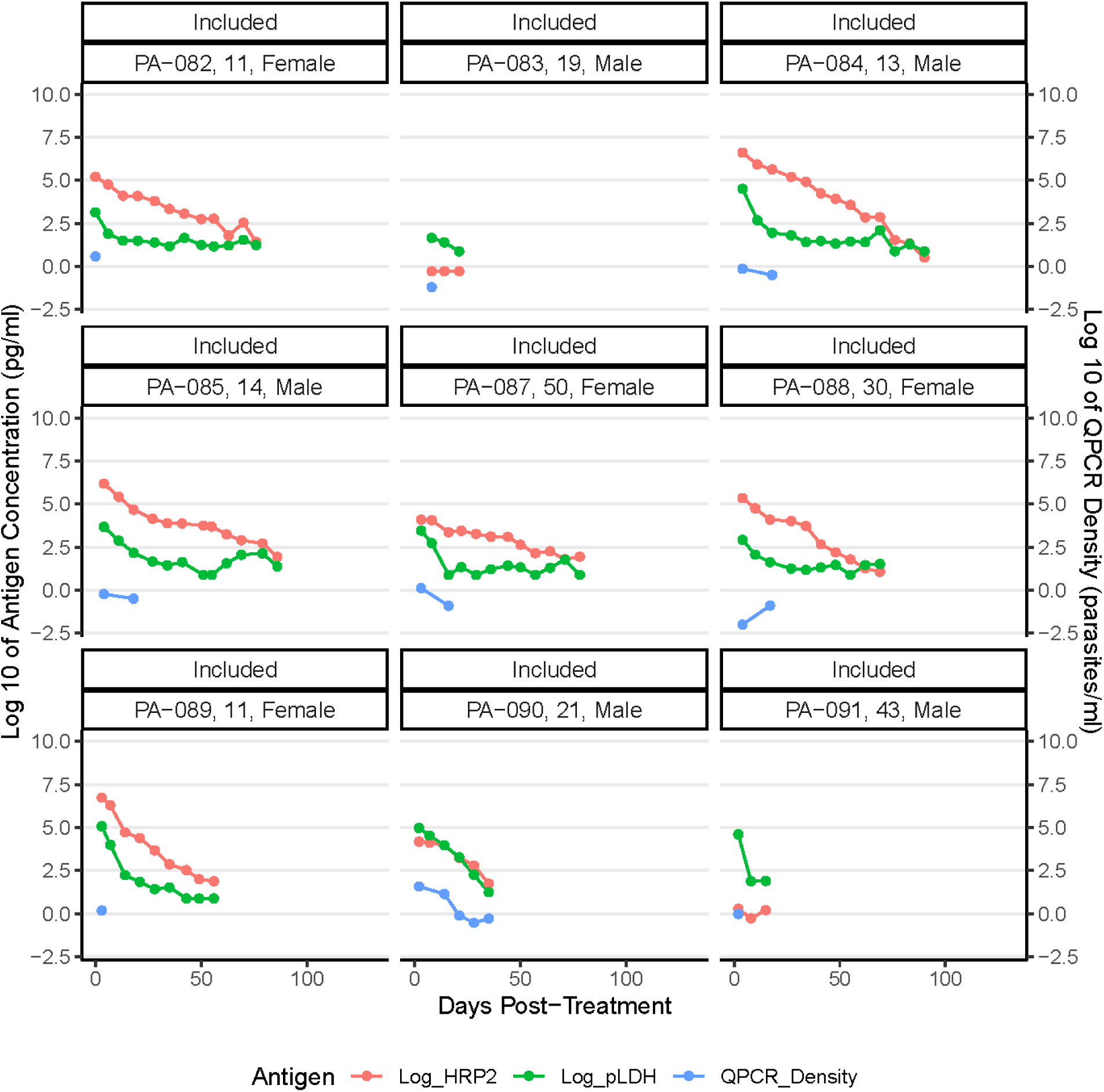
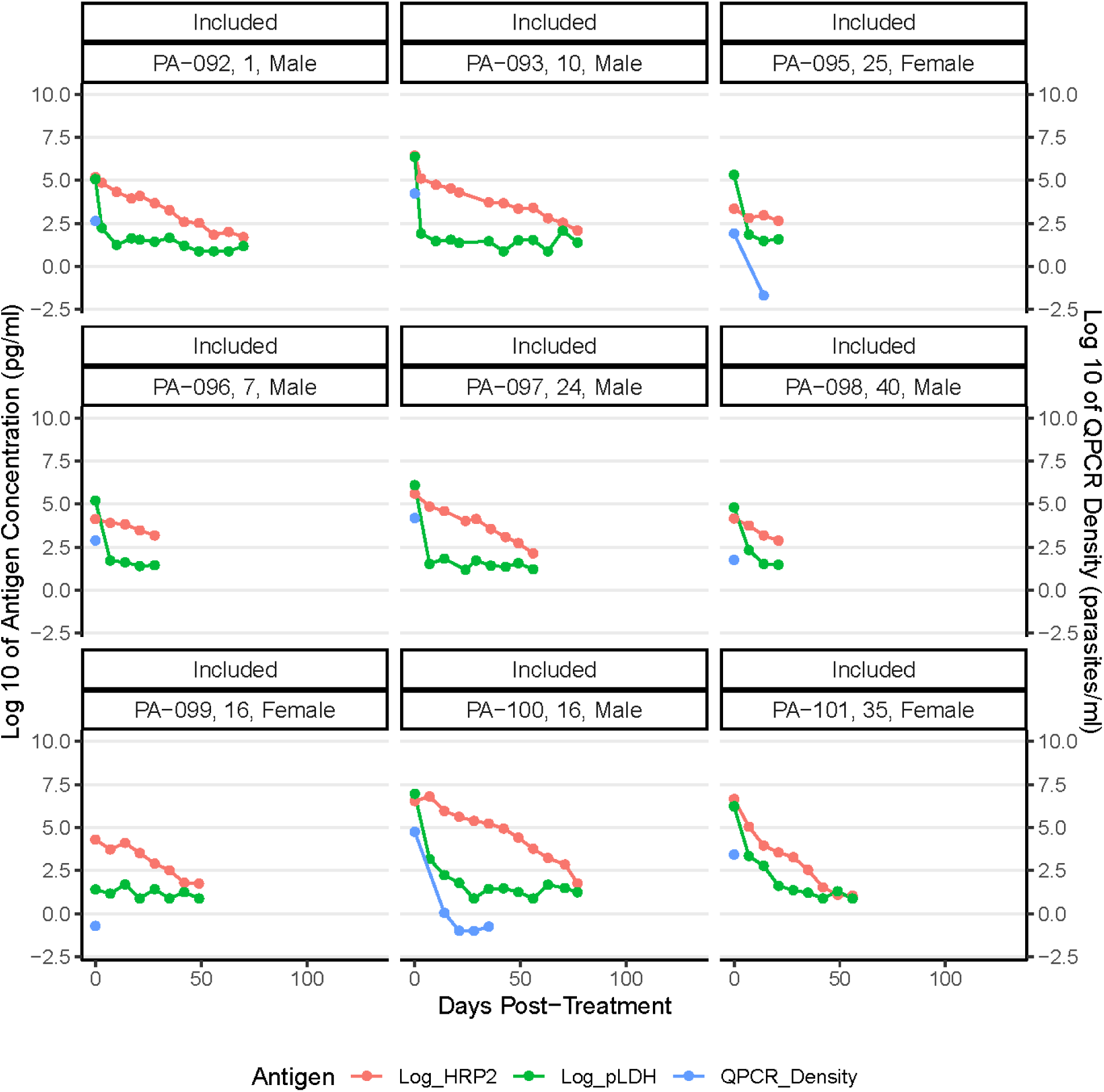
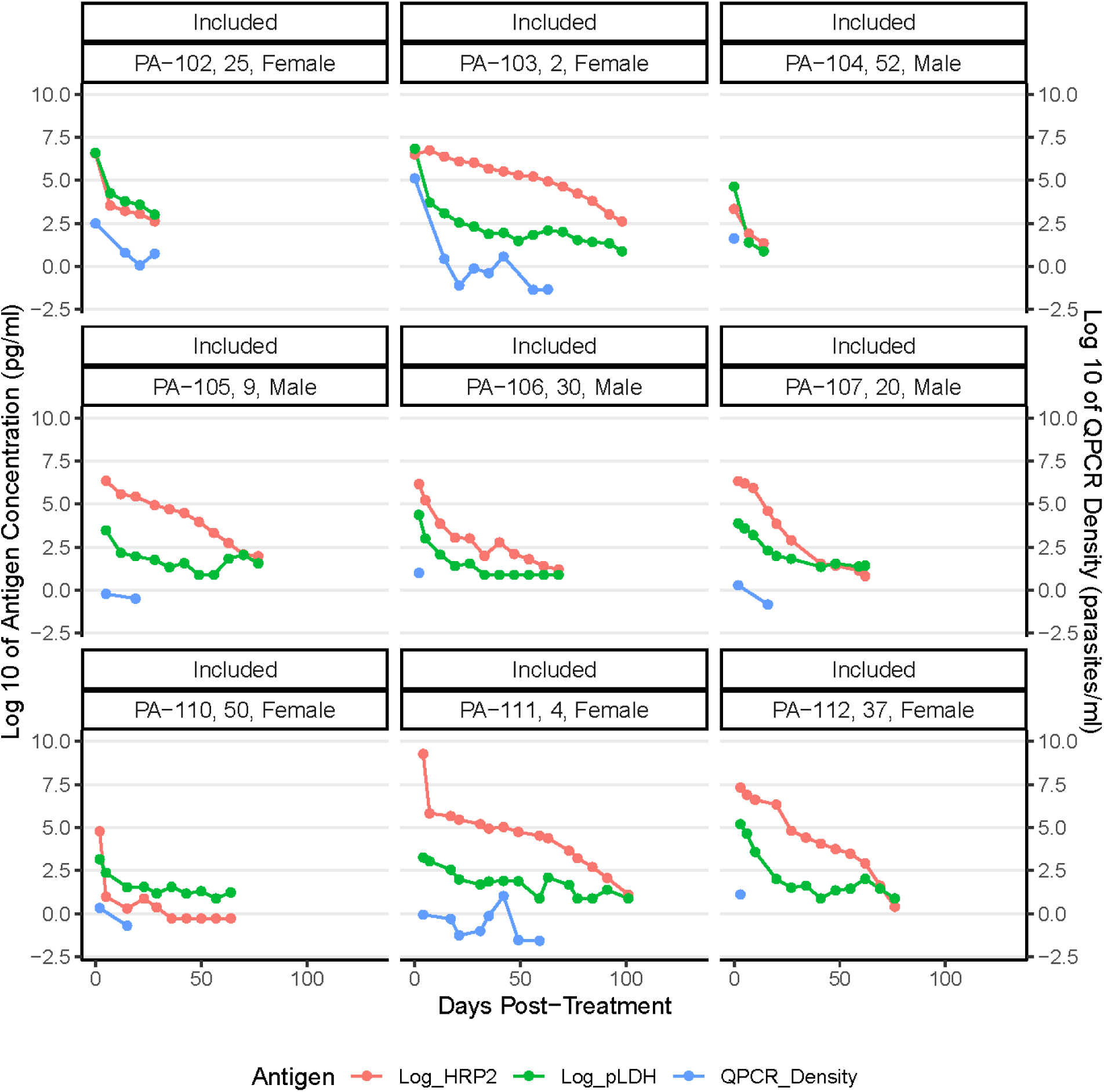
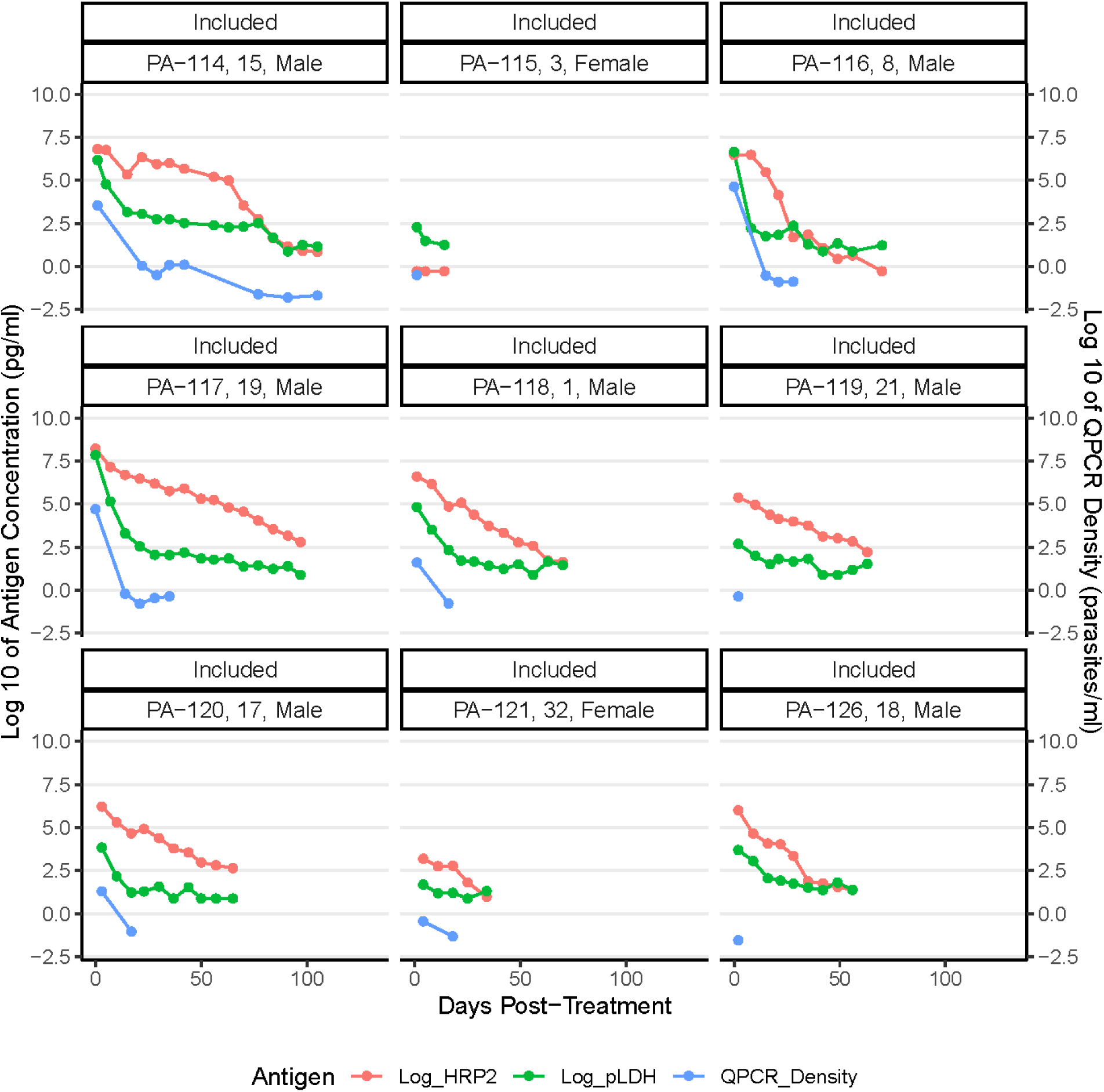
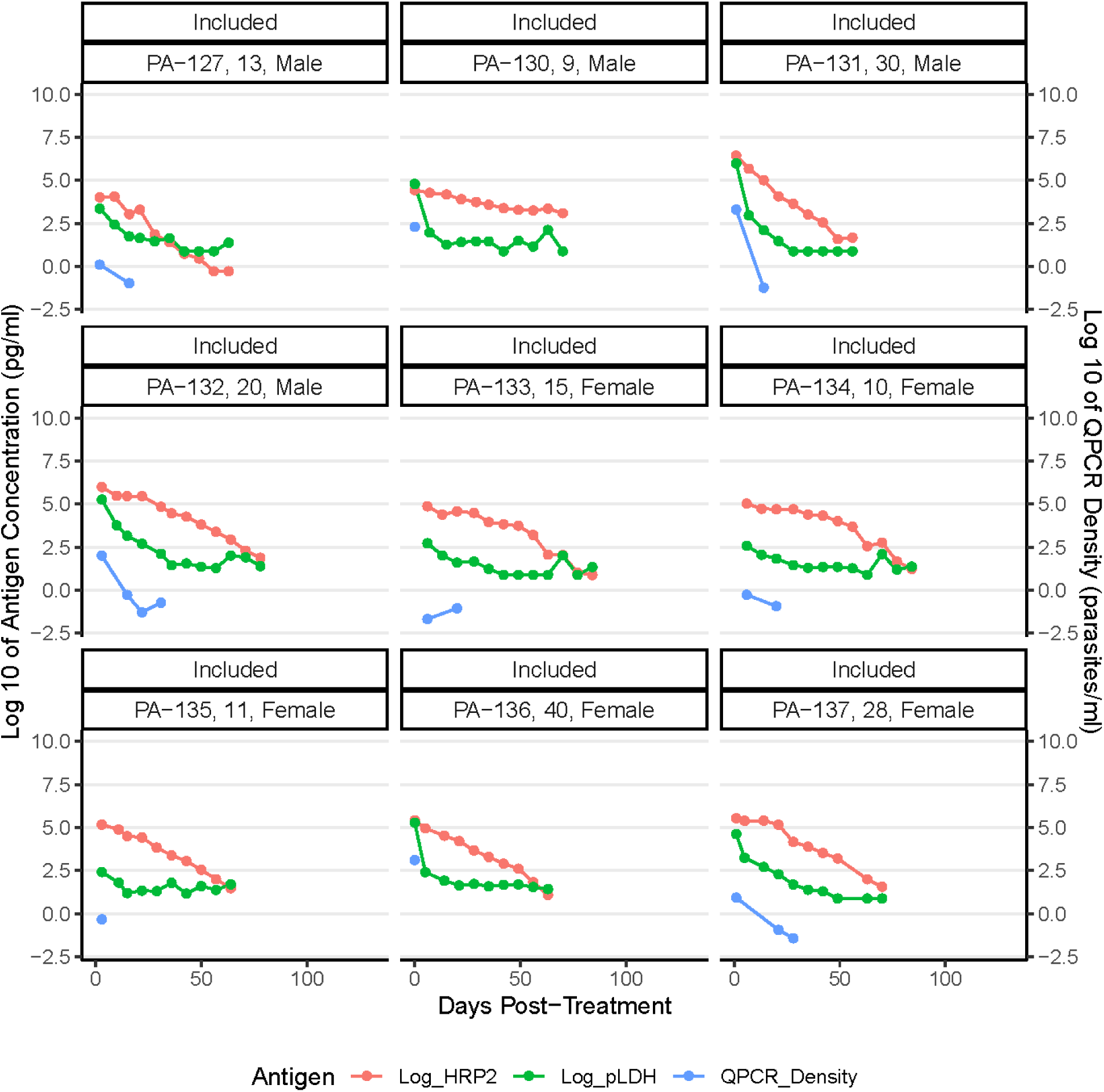
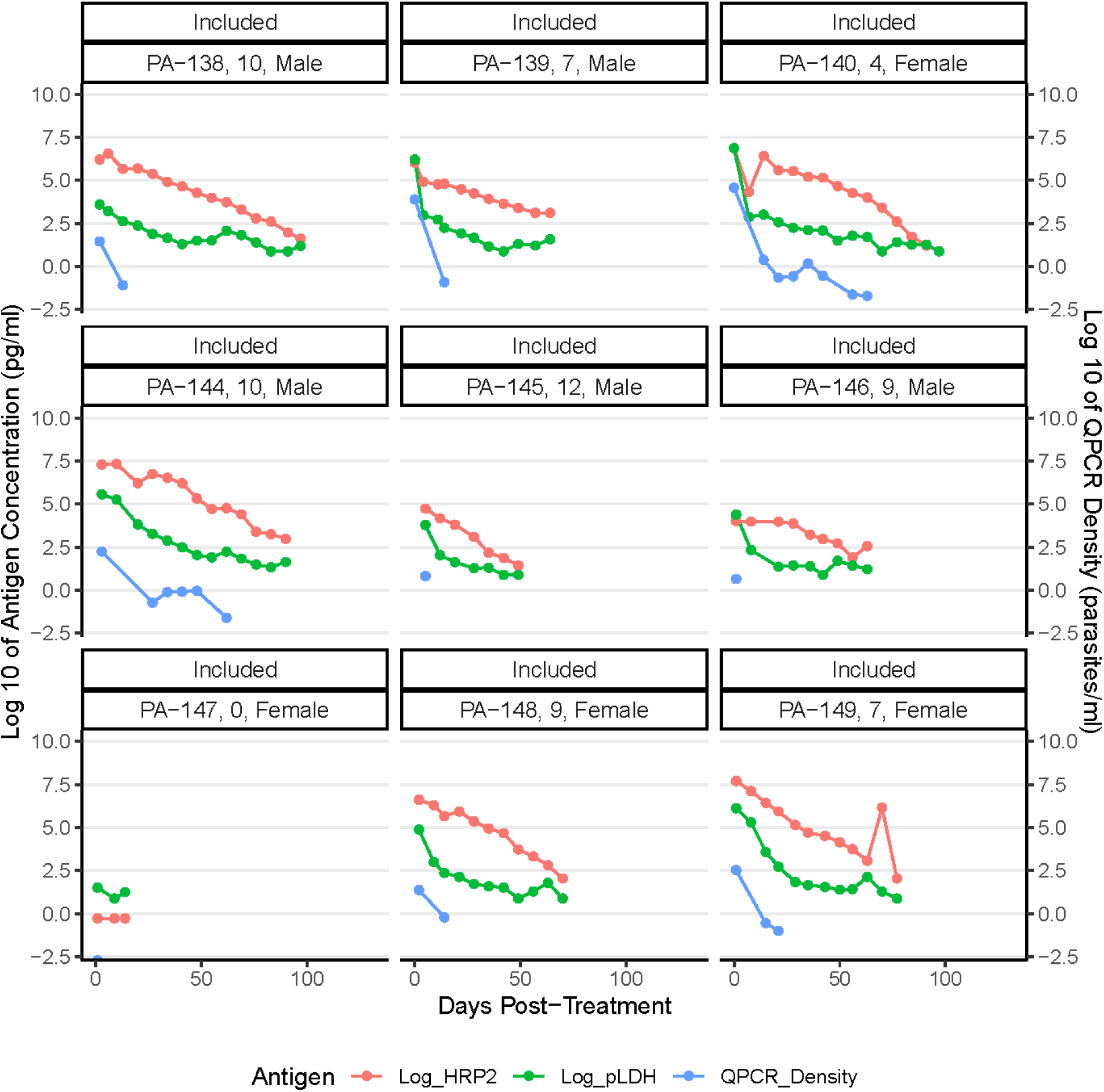
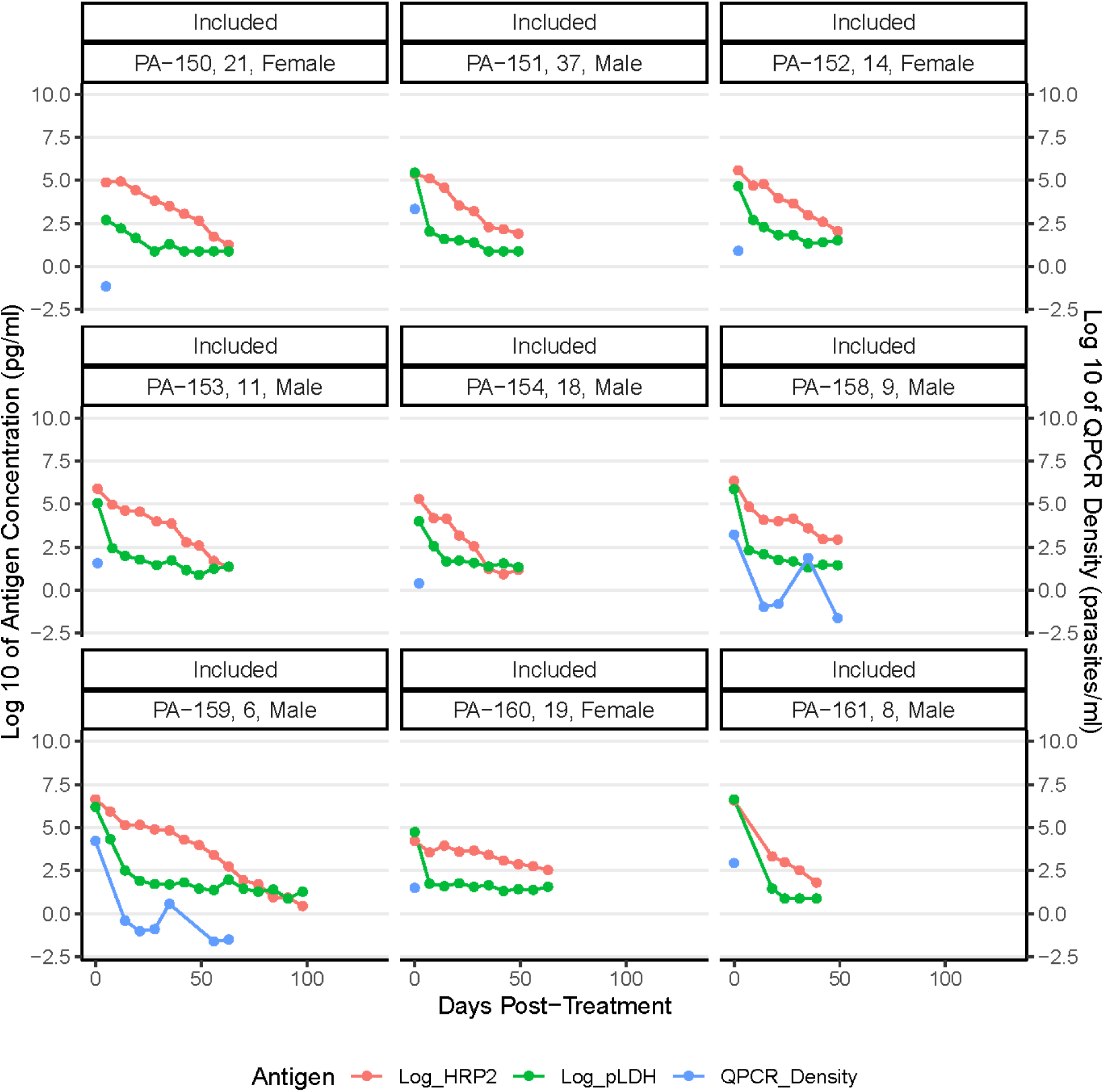
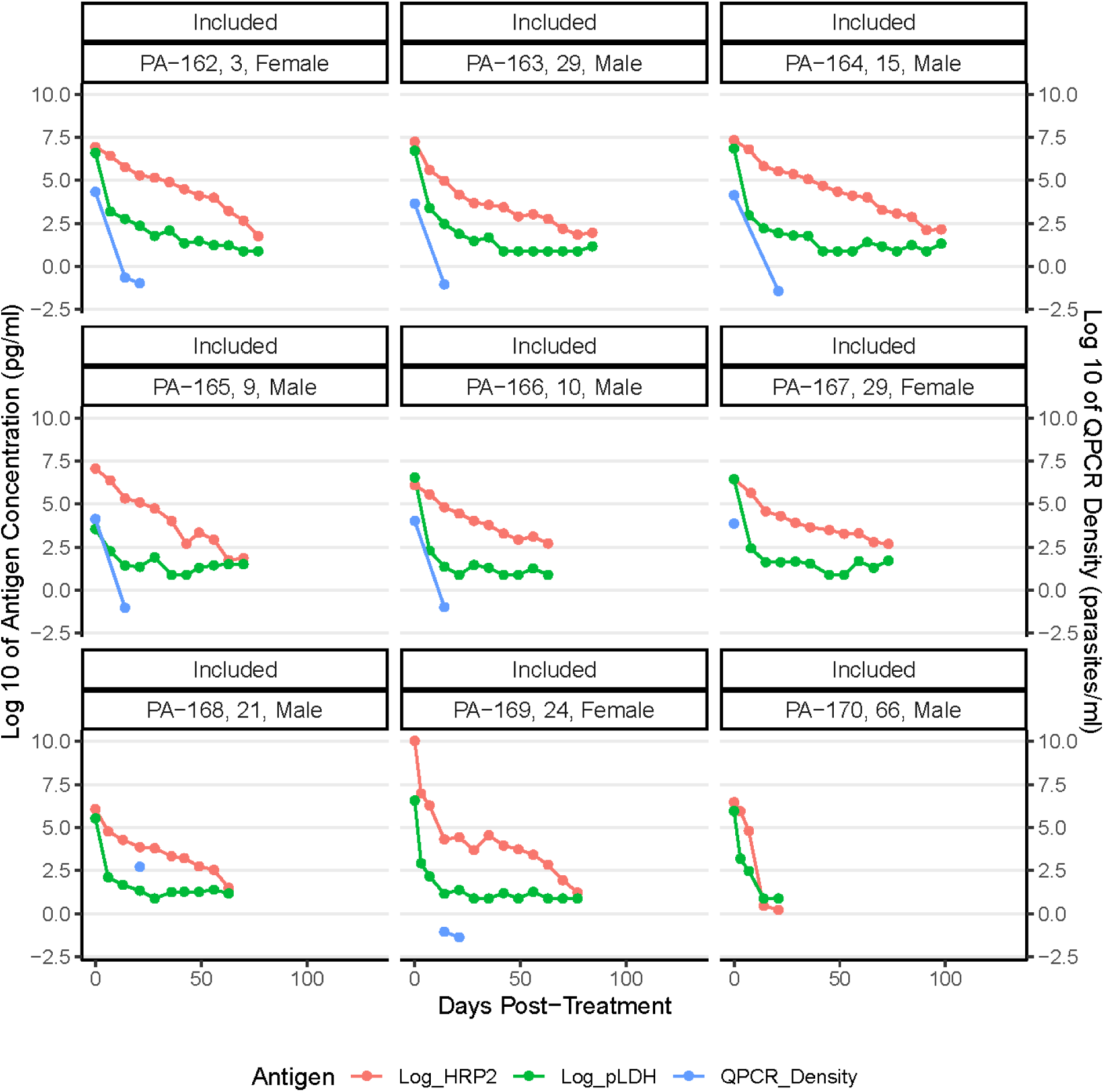
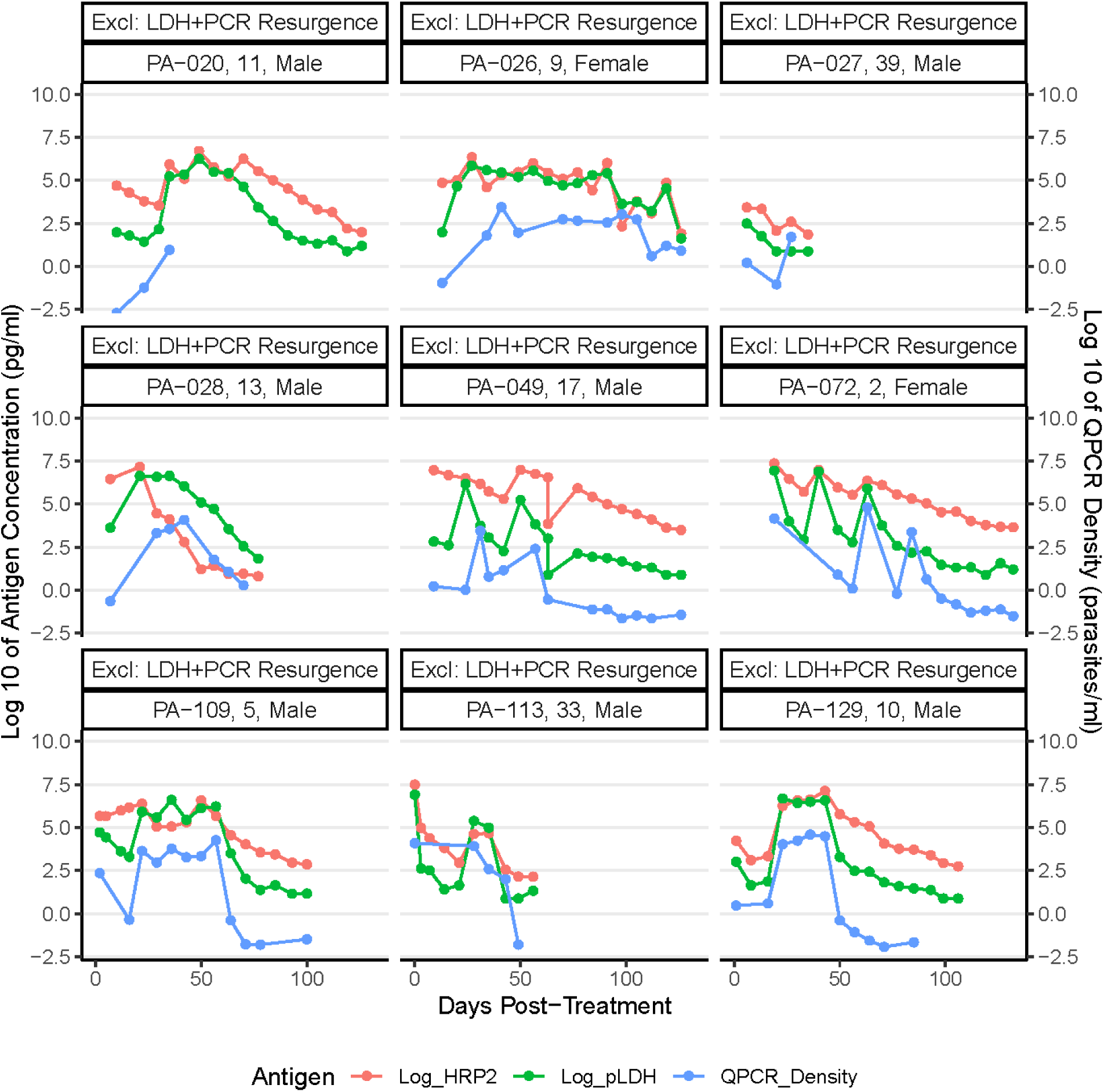
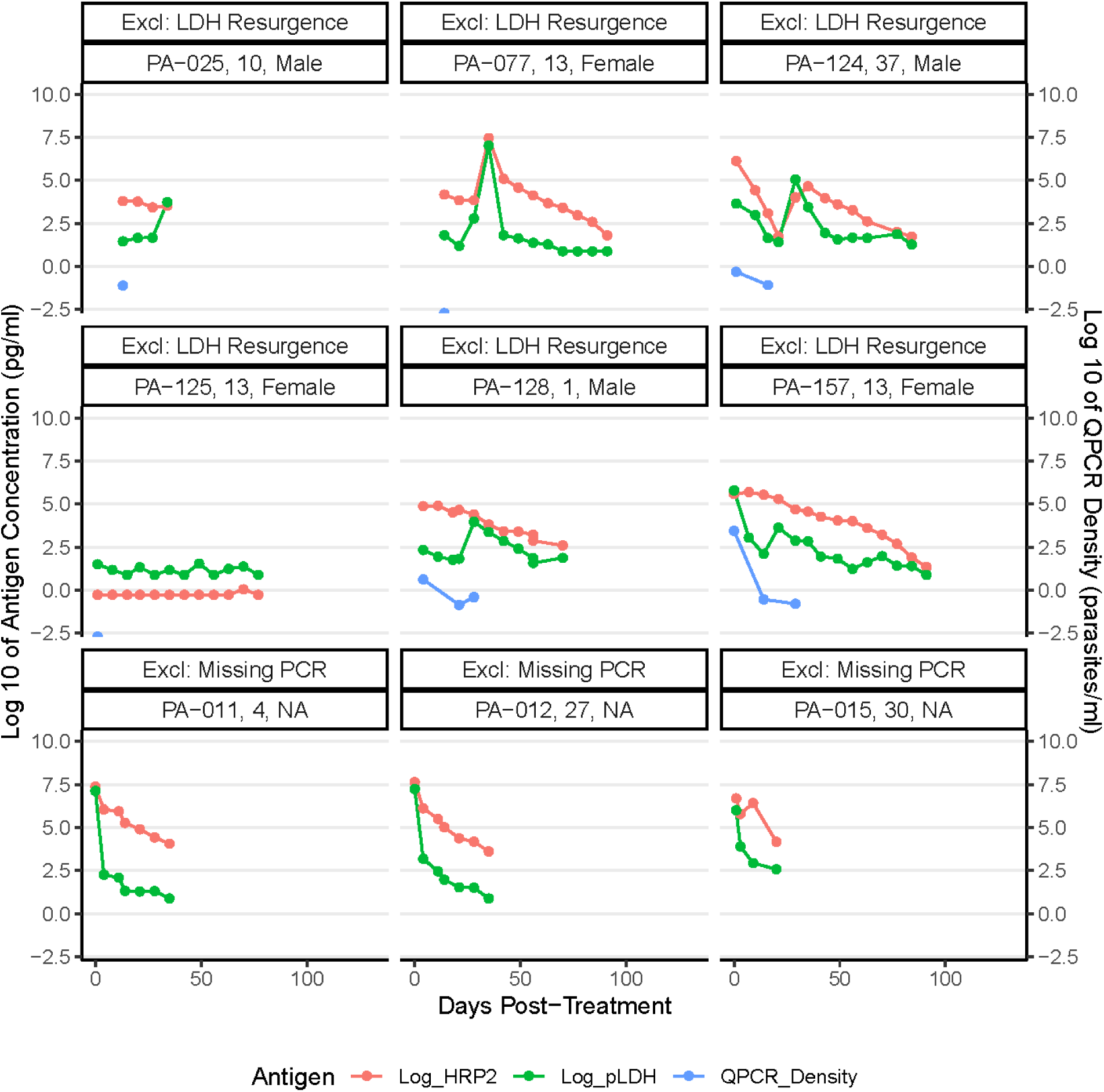
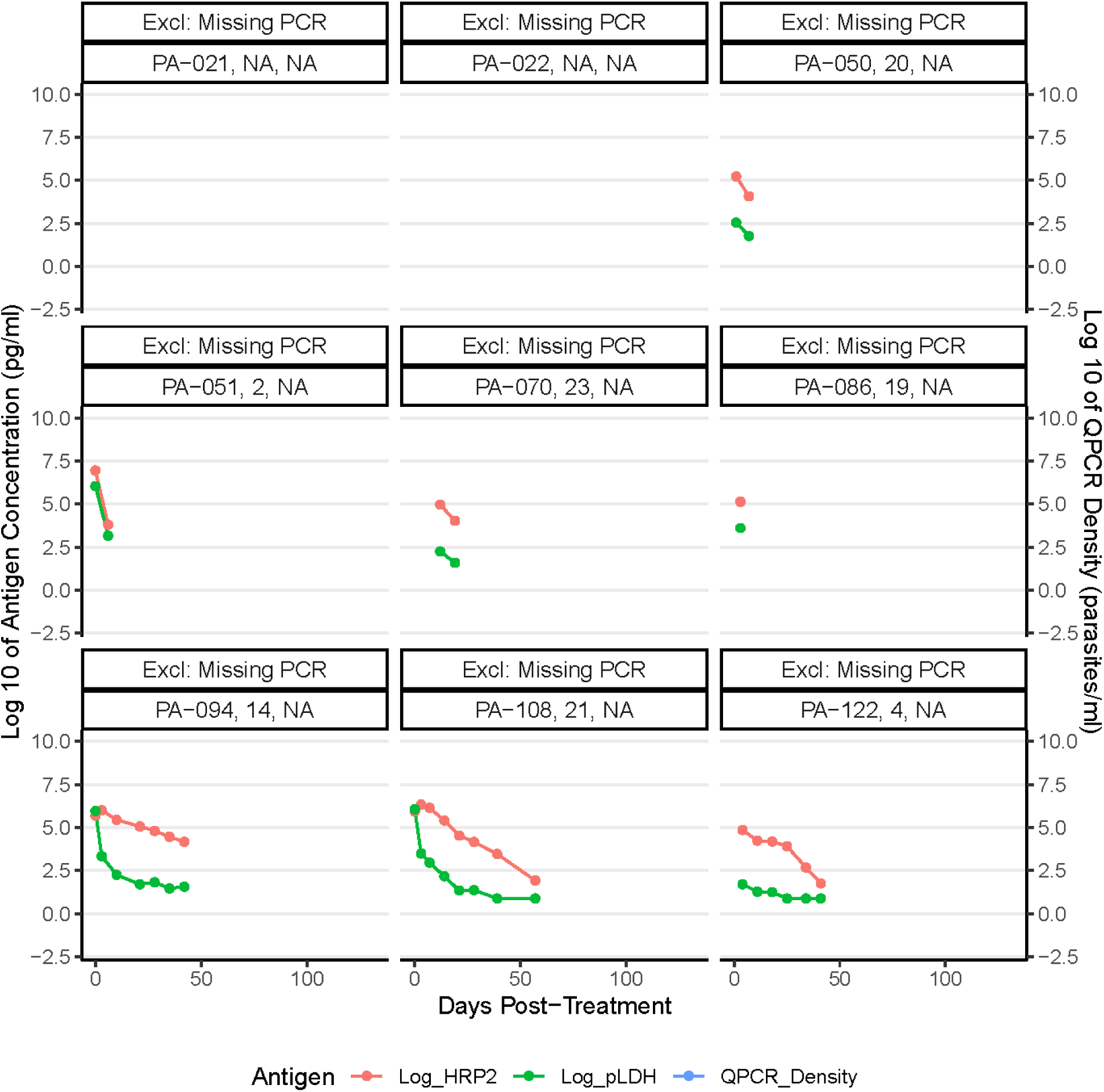
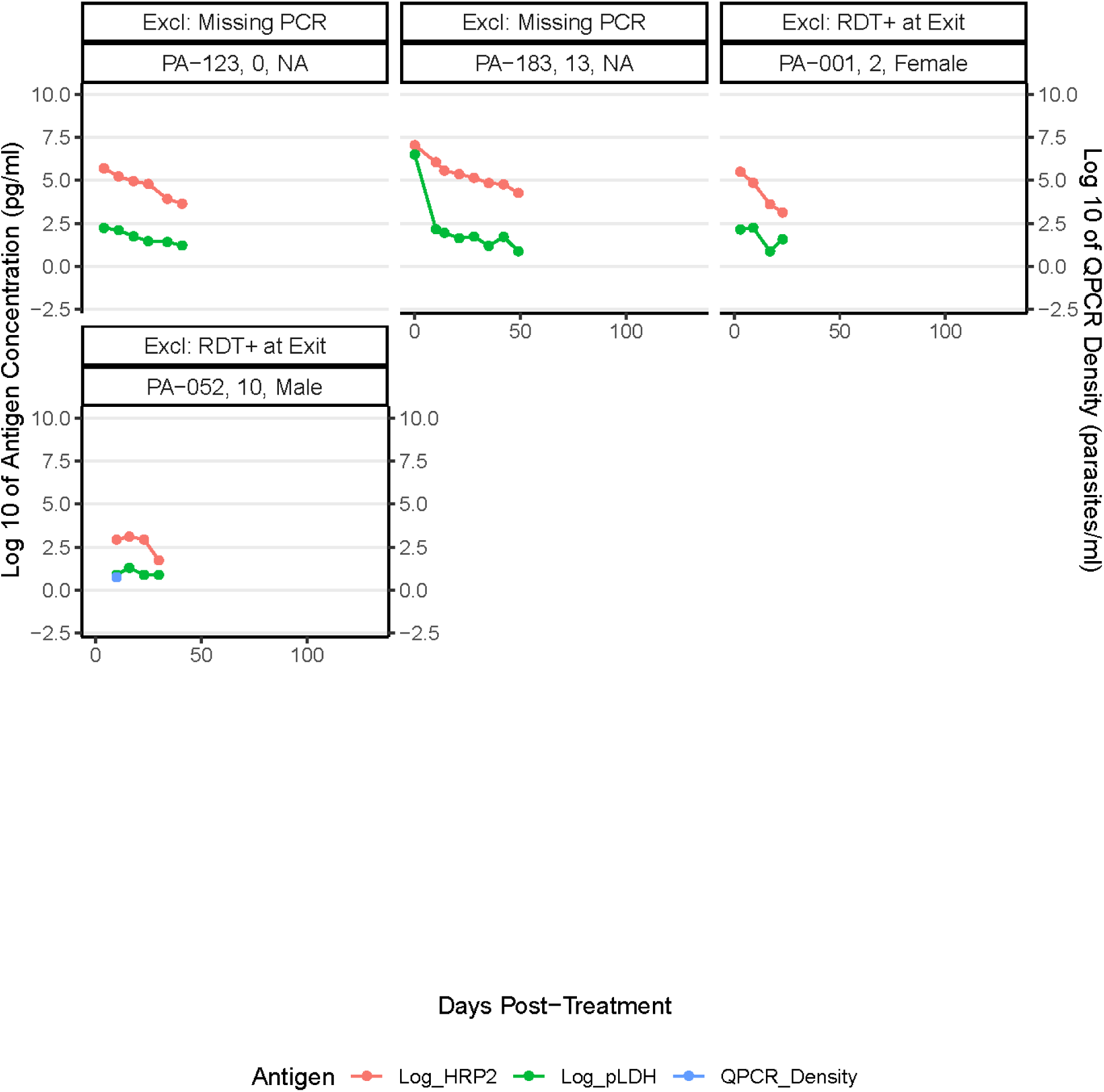
Individual participant antigen dynamic profiles showing the log-transformed HRP2 and PfLDH concentrations for each participant over time, as well as QPCR density where available. Each participant has their ID number, age, sex, and exclusion criteria labeled.

**Supplementary Figure 3:**
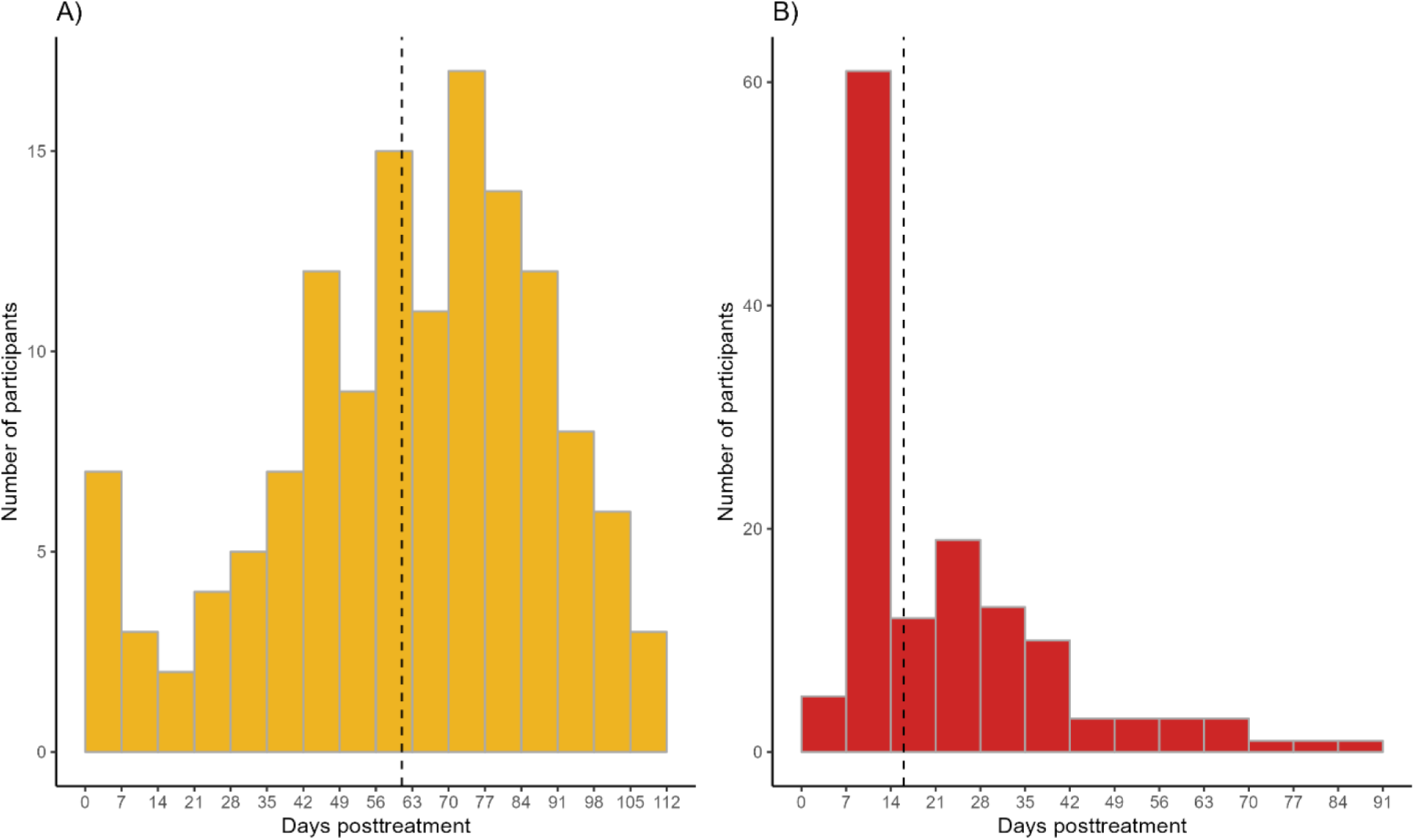
Histogram of the time post-treatment to reach a threshold of 80 pg/ml for each antigen. HRP2 is shown in panel A on the left, pLDH is shown in panel B on the right. The dashed lines indicate the mean time to reach 80 pg/ml for each antigen.

**Supplementary Table 1:**
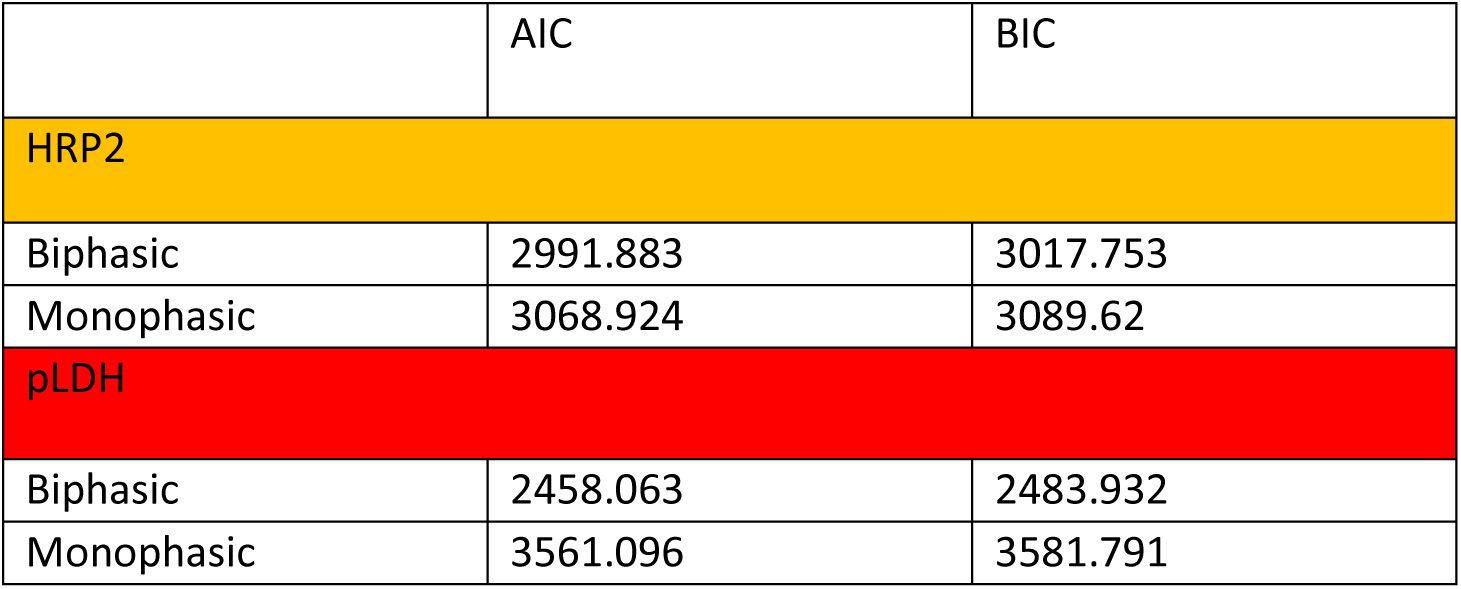
Table displaying AIC and BIC for the biphasic and monophasic models respectively for each target antigen.

|  | AIC | BIC |
| --- | --- | --- |
| **HRP2** |  |  |
| Biphasic | 2991.883 | 3017.753 |
| Monophasic | 3068.924 | 3089.62 |
| **pLDH** |  |  |
| Biphasic | 2458.063 | 2483.932 |
| Monophasic | 3561.096 | 3581.791 |

**Supplementary Table 2:** Table displaying HRP2 and PfLDH antigen concentrations identified at 90% probability of positivity for Rapigen BIOCREDIT and WHO PQ comparator RDTs. Adapted from Golden et al., 2023.^16^

| Test | Antigen concentration at which test has 90% probability of positivity |  |  |  |
| --- | --- | --- | --- | --- |
|  | HRP2 (pg/mL) |  | PfLDH (pg/mL) |  |
|  | Median estimate | 95% credible interval | Median estimate | 95% credible interval |
| Rapigen Pf (pLDH/HRPII) | 525 | (407-661) | 1,318 | (1,175-1,479) |
| Rapigen Pf/Pv (pLDH/pLDH) | - | - | 372 | (302-477) |
| WHO PQ comparator HRP2/PfLDH RDT | 1,072 | (955-1,202) | 5,754 | (5,012-6,607) |

